# Machine learning models to identify patient and microbial genetic factors associated with carbapenem-resistant *Klebsiella pneumoniae* infection

**DOI:** 10.1101/2020.07.06.20147306

**Authors:** Zena Lapp, Jennifer H Han, Jenna Wiens, Ellie JC Goldstein, Ebbing Lautenbach, Evan S Snitkin

**Author notes:** Corresponding author: Evan Snitkin, Address: 1520D MSRB I, 1150 W. Medical Center Dr., Ann Arbor, MI, 48109-5680.

## Abstract

Carbapenem-resistant *Klebsiella pneumoniae* (CRKP) is a critical-priority antibiotic resistance threat that has emerged over the past several decades, spread across the globe, and accumulated resistance to last-line antibiotic agents. While CRKP infections are associated with high mortality, only a small subset of patients acquiring CRKP colonization will develop clinical infection. Here, we sought to determine the relative importance of patient characteristics and CRKP genetic background in determining patient risk of infection. Machine learning models classifying colonization vs. infection were built using whole-genome sequences and clinical metadata from a comprehensive set of 331 CRKP isolates collected across 21 long-term acute care hospitals over the course of a year. Model performance was evaluated based on area under the receiver operating characteristics curve (AUROC) on held-out test data. We found that patient and genomic features were predictive of clinical CRKP infection to similar extents (AUROC IQRs: patient=0.59-0.68, genomic=0.55-0.61, combined=0.62-0.68). Patient predictors of infection included the presence of indwelling devices, kidney disease and length of stay. Genomic predictors of infection included presence of the ICEKp10 mobile genetic element carrying the yersiniabactin iron acquisition system, and disruption of an O-antigen biosynthetic gene in a sub-lineage of the epidemic ST258 clone. Altered O-antigen biosynthesis increased association with the respiratory tract, and subsequent ICEKp10 acquisition was associated with increased virulence. These results highlight the potential of integrated models including both patient and microbial features to provide a more holistic understanding of patient clinical trajectories.

**Importance:** Multidrug resistant organisms, such as carbapenem-resistant Klebsiella pneumoniae (CRKP), colonize alarmingly large fractions of patients in endemic regions, but only a subset of patients develop life-threatening infections. While patient characteristics influence risk for infection, the relative contribution of microbial genetic background to patient risk remains unclear. We used machine learning to determine whether patient and/or microbial characteristics can discriminate between CRKP colonization vs. infection across multiple healthcare facilities and found that both patient and microbial factors were predictive. Examination of informative microbial genetic features revealed features associated with respiratory colonization and higher rates of infection. The methods and findings presented here provide a foundation for future epidemiological, clinical, and biological studies to better understand bacterial infections and clinical outcomes.

## Introduction

Infections due to multidrug resistant organisms lead to hundreds of thousands of deaths worldwide each year (1). Carbapenem-resistant Enterobacterales (CRE) are a critical-priority antibiotic resistance threat that has emerged over the past several decades, spread across the globe, and accumulated resistance to last-line antibiotic agents (2, 3). In the United States (U.S.), CRE infections are primarily caused by the sequence type (ST) 258 strain of carbapenem resistant *Klebsiella pneumoniae* (CRKP), which has become endemic in regional healthcare networks (3–7). In this background of regional endemicity the risk of patient exposure to CRKP is high, as evidenced by alarmingly high rates of colonization, especially in long-term care settings (7, 8). However, even among critically ill patients residing in long-term care facilities, not all colonized patients develop clinical infections that require antibiotic treatment (3, 9). Currently, our understanding of the factors that influence whether a colonized patient develops an infection is incomplete.

In addition to clinical characteristics of patients, the genetic background of the colonizing strain may also influence the risk of infection, as there is extensive intra-species variation in antibiotic resistance and virulence determinants harbored by *K. pneumoniae* (3). To date, most studies of virulence determinants have been carried out in model systems (10, 11) or examined in human populations without considering patient characteristics or clinical context (11, 12). One recent study investigated virulence determinants in *K. pneumoniae* clinical isolates while controlling for patient characteristics (13). However, this was a single-site study with a focus on carbapenem-susceptible *K. pneumoniae*, thereby not addressing the impact of genomic variation in antibiotic-resistant lineages that circulate in global healthcare systems.

Here, we sought to understand the importance of both patient factors and CRKP genetic background in determining whether a patient is infected (vs. colonized) with CRKP, and identify a set of patient and microbial features that are consistent predictors of CRKP infection across long-term care facilities. To accomplish this, we compared patients with CRKP colonization and infection based upon both clinical characteristics and the genomes of their colonizing or infecting strains. We leveraged a comprehensive set of all clinical isolates collected from 21 long-term acute care hospitals (LTACHs) across the U.S. over the course of a year to obtain an unbiased assessment of the relative contribution of bacterial and host factors to patient colonization and infection with circulating CRKP lineages. To do this, we employed a rigorous machine learning approach that allowed us to perform a direct comparison of patient and bacterial features, as well as identify the most predictive features across our unbiased study population.

## Results

Of 355 clinical CRKP isolates from 21 LTACHs across the U.S. (14), we classified 149 (42%) of the isolates as representing infection based on modified NHSN criteria (**Figure S2, Tables S1-3**). Stratified by anatomic site, we classified 29/29 (100%) blood isolates as infection, 69/196 (35%) respiratory isolates as infection, and 51/130 (39%) urinary isolates as infection (**Table S3**). More than 90% of patient isolates were from the epidemic CRKP lineage ST258 (**Tables S1**). Patients harboring different sequence types of CRKP showed no significant differences in infection/colonization status or anatomic site of isolation, and no substantive differences in clinical characteristics (see supplementary material). Thus, we decided to limit our analysis to ST258 to improve our ability to discern whether genetic variation within this dominant strain is associated with infection.

### The CRKP epidemic lineage ST258 shows evidence of sub-lineage variation in virulence and anatomic site of isolation

We next evaluated if there exist sub-lineages of ST258 with altered virulence properties by looking for clustering of isolates by infection on the whole-genome phylogeny (**Figure 1**; see supplementary methods) (38). Infection status was non-randomly distributed on the phylogeny (p=0.002), supporting our hypothesis that the genetic background of CRKP influences infection. We performed a similar clustering analysis to look at potential niche-specific adaptation to certain anatomic sites (**Figure 1**), and found that respiratory (p=0.001) and urinary (p=0.013) isolates cluster on the phylogeny, but blood isolates do not (p=0.21). This analysis indicates that, in addition to patient features, intra-strain variation in virulence and adaptation to the urinary and respiratory tract might influence whether patients develop an infection.

**Figure 1:**
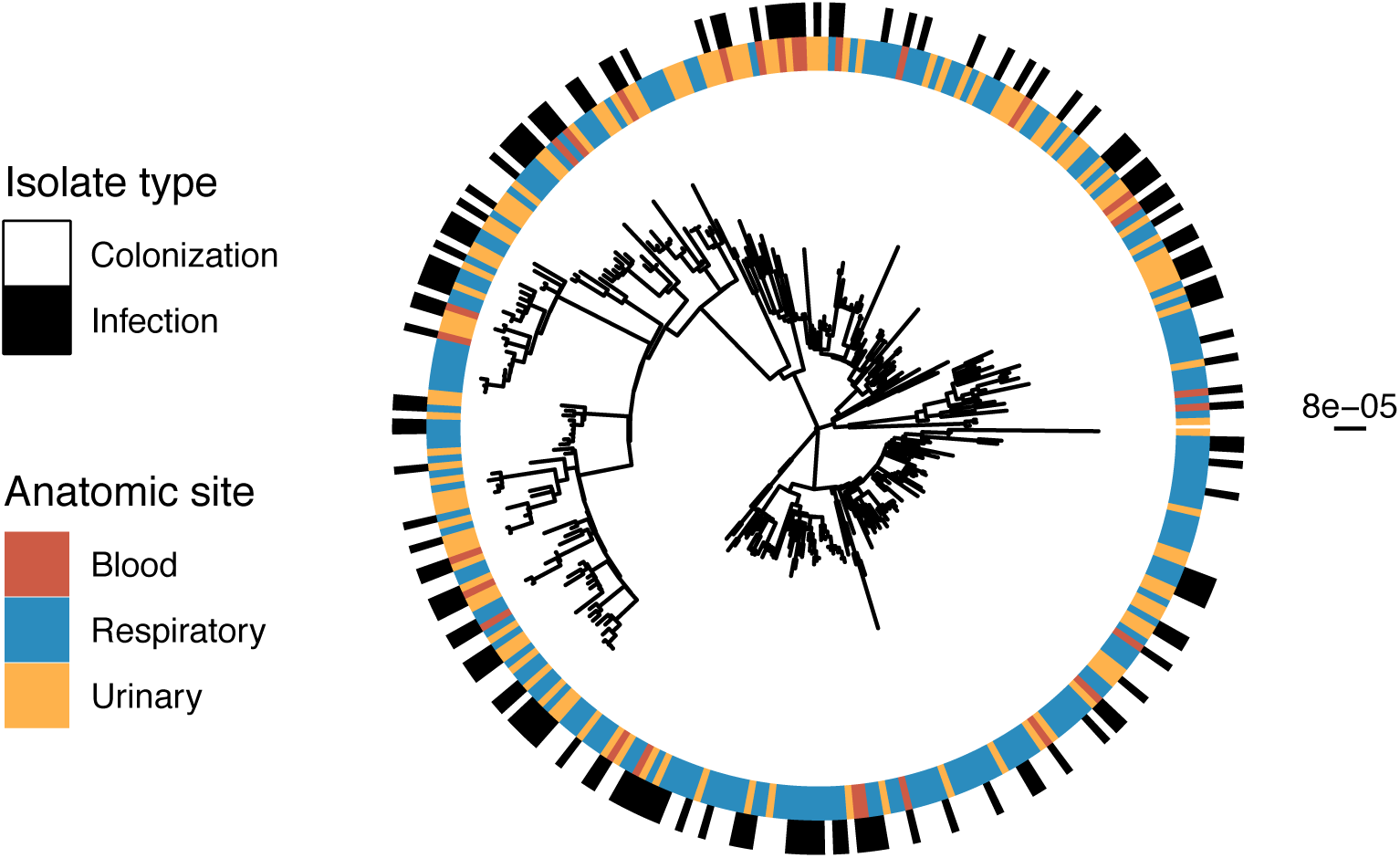
Infection and anatomic site cluster on the phylogeny. Maximum likelihood phylogenetic tree of all isolates including infection or colonization classification for each isolate and anatomic site of isolation. The scale bar to the right of the phylogeny shows the branch length in substitutions per site. Testing for non-random distribution of isolates on the phylogeny (see supplementary methods) revealed clustering of infection, respiratory, and urinary isolates on the phylogeny, respectively.

### Both patient and CRKP genetic characteristics are weakly predictive of infection, with relative performance being highly facility-dependent

We next performed machine learning to quantify the ability of patient and microbial genetic characteristics to predict CRKP infection (**Figure S1**). To prevent over- or under-fitting and control for facility-level biases, we generated 100 train/test data splits, wherein a given LTACH was only included either in the train or test set. Each LTACH occurred a median of 24 times (range 13-32) in the test data split. In this way, we were able to identify patient and CRKP strain characteristics consistently associated with infection or colonization across data splits, and thus across patient populations in different healthcare facilities.

First, we sought to understand if patient and genomic features were individually predictive of CRKP infection. To this end, we independently evaluated patient characteristics as well as three different genomic feature sets for their ability to classify colonization and infection. The three genomic feature sets were uncurated genomic (including SNPs, indels, IS elements, and accessory genes), uncurated grouped genomic (variants grouped into genes, akin to a burden test, e.g (28)), and curated genomic (features identified using Kleborate (16)). Across the 100 different train/test splits, we observed that the average predictive performance was weak, with each of the genomic and patient feature sets predictive of infection to a similar degree (all 1st quartile AUROCs > 0.5; median range=0.55-0.68; **Figure 2A**; AUPRC: **Figure S3A**).

**Figure 2:**
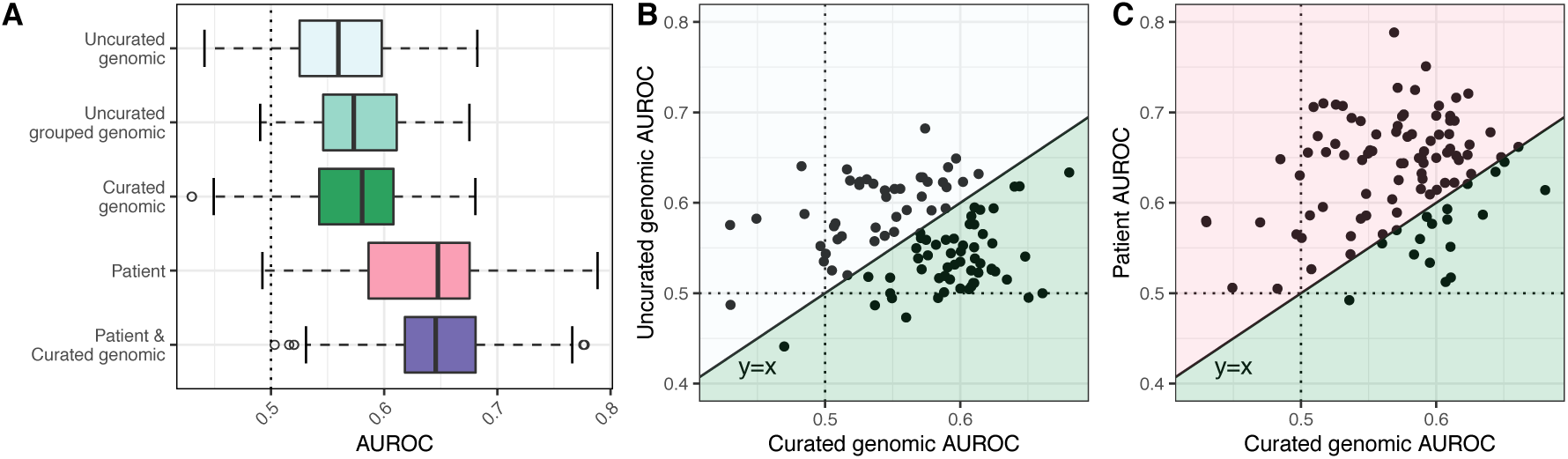
Test AUROCs for various classifiers identifying CRKP colonization vs. infection vary substantially across data splits. (A) Test AUROCs for 100 L2 regularized logistic regression models different using train/test splits. All isolates from a given LTACH were included in either the training split or the testing split for each data split. We built models using five different feature sets, keeping the same 100 data splits. AUROCs of different feature sets were not significantly different. In the right two panels, the curated genomic feature set AUROCs are compared to: (B) the uncurated genomic feature set AUROCs, and (C) the patient feature set AUROCs. Each point is the resulting pair of AUROCs for models built with the same data split, but the two respective feature sets. The dotted lines in all 3 panels indicate the AUROC for choosing an outcome randomly (0.5); anything below the line is worse than random, and anything above the line is better than random. The solid diagonal line in the right two panels is the line y=x; points below the line correspond to a higher curated genomic AUROC for that data split, and points above the line correspond to a higher uncurated genomic AUROC (B), or patient AUROC (C), respectively. The colors in panels (B) and (C) correspond to the colors in panel (A); the points in a given colored area indicate that that feature set had the higher AUROC for that data split. In both cases, one feature set does not consistently outperform the other (p=0.4; see supplementary methods for p-value calculation). AUROC=area under the receiver operating characteristic curve.

Additionally, no one feature set was consistently the most predictive (e.g. **Figures 2B, 2C**; all comparisons p > 0.30, see supplementary methods for p-value calculation). Furthermore, for each feature set the AUROCs were distributed such that the test AUROC ranged from below 0.5 to over 0.7, depending on how the data were split (i.e., which facilities appear in the train/test sets). This variation in model performance across different train/test sets suggests that the association of CRKP strain and patient characteristics with infection or colonization varies across facilities.

### Integration of patient and CRKP strain features does not improve discriminative performance of overall or anatomic site-specific models

To determine if the predictive power of patient and genomic features is additive, and if combining these disparate feature sets improved validation on held-out facilities, we built models including both patient and curated genomic features. The discriminative performance of the models based on the combined feature set was not significantly greater than that of the individual feature sets (**Figure 2A**, p ≥ 0.20). Thus, despite variation in the predictive capacity of genomic and patient features across facilities (**Figure 2C**), combining the two sets did not improve overall performance. Focusing on anatomic site-specific models revealed similar trends, where classification performances were similar for respiratory and urinary specific models, and the relative predictive capacity of patient and CRKP strain features varied across facility subsets (**Figure S4**; AUPRC: **Figure S3B**).

### Some patient and genomic features consistently discriminate colonization and infection

After evaluating the predictive capacity of models, we next sought to identify patient and CRKP strain characteristics that are most associated with CRKP infection or colonization. To this end, we identified those patient and genomic features that consistently improved model performance across the 100 different data splits (see methods). Evaluating the importance of features in this way provides insight into those characteristics that generalize across different facility subsets. This approach was taken for both overall and anatomic site-specific models to identify features predictive of different anatomic sites of infection (**Figure 3, Figures S5-7**).

**Figure 3:**
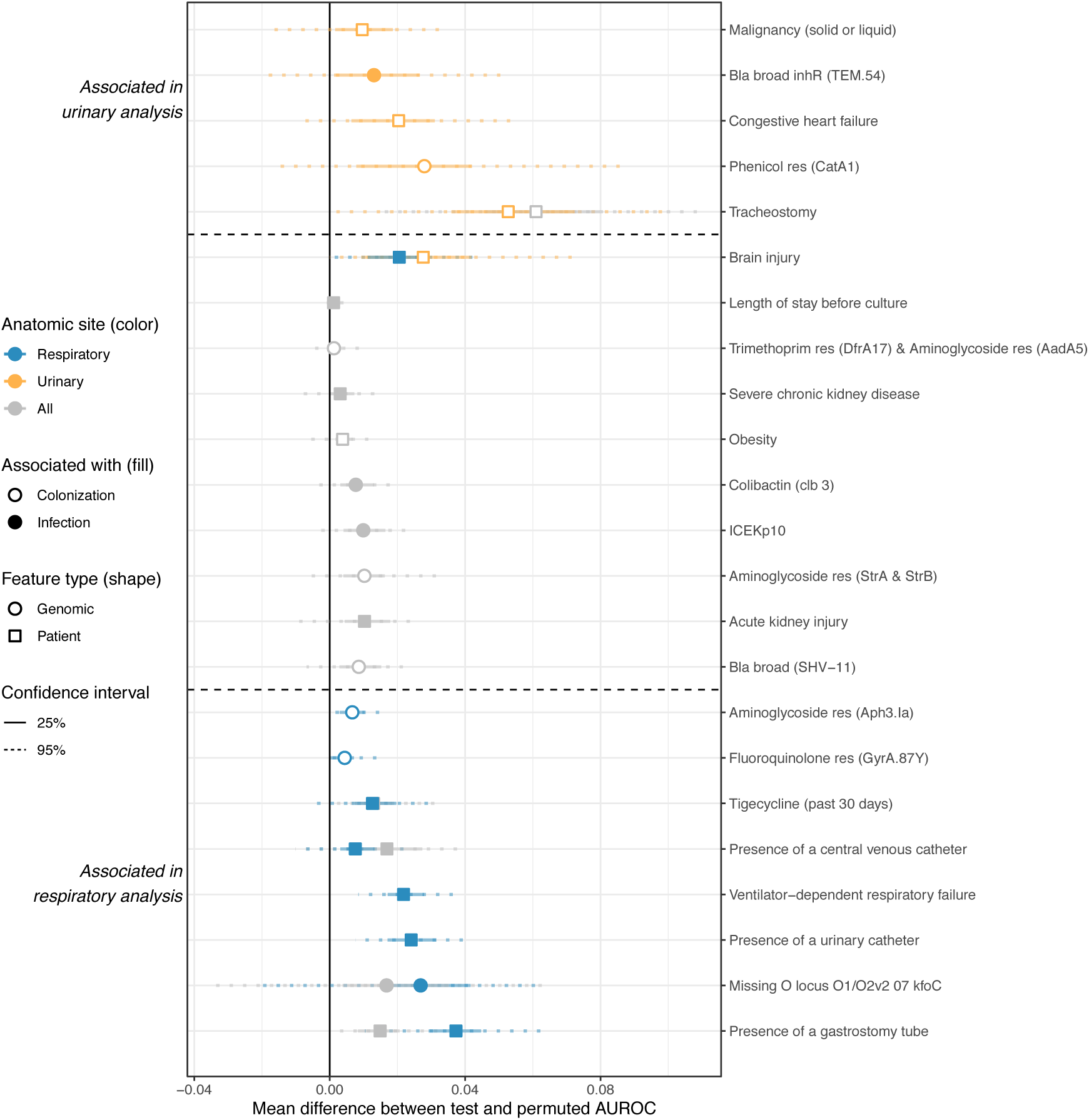
Features consistently associated with colonization or infection sometimes differ between the overall, respiratory, and urinary models. Feature-specific improvement in model performance, measured as the mean difference between test and permuted AUROC (see methods), of features found to be consistently associated with colonization or infection in at least one of the following analyses: overall, respiratory-specific, urinary-specific. We consider features to be associated with infection/colonization if the AUROC difference was greater than zero in over 75% of the 100 data splits. The vertical solid black line indicates a difference of zero (i.e. the feature provides no improvement to model performance). Horizontal dotted lines separate features associated with urinary but not respiratory isolates (top), both urinary and respiratory (or all) isolates (middle), or respiratory but not urinary isolates (bottom). Bla=Beta lactamase, res=confers resistance to that antibiotic class. AUROC=area under the receiver operating characteristic curve.

Several patient features were consistently associated with infection in the overall analysis, including presence of a gastrostomy tube, presence of a central venous catheter, acute kidney injury, and severe chronic kidney disease (**Figure 3**), all markers of critically ill patients. Only a small number of genomic features were consistently associated with infection (**Figure 3**). No known virulence factors were positively associated with colonization; all of the genomic features positively associated with colonization are antibiotic resistance elements. Conversely, all but one of the genomic features positively associated with infection (3/4) are related to virulence. The ICEKp10 element is positively associated with infection and carries colibactin and two different types of yersiniabactin, a previously identified *K. pneumoniae* virulence determinant (12) (**Figure S8**). Additionally, insertion sequence-mediated disruption of the O-antigen biosynthetic gene *kfoC* (see supplementary methods and **Figure S9** for insertion sequence identification (21, 23, 39–41)) was associated with respiratory infection. Colibactin is a toxin (3), and yersiniabactin is an iron scavenging system that has been identified in previous animal and human studies as being associated with virulence (3, 10). The O-antigen of lipopolysaccharide (LPS) is a known antigenic marker, although association with a specific anatomic site has not been noted (42).

### A sub-lineage of ST258 clade II appears to have sequentially evolved enhanced adaptation for the respiratory tract and increased virulence

We noted that *kfoC* disruption is largely confined to a sub-lineage of ST258 present across 12 LTACHs in California (**Figures 4, S10, S11**). Consistent with this feature being associated with respiratory infection, the disrupted *kfoC* lineage is enriched in respiratory isolates (82/118, 69% of isolates in the disrupted *kfoC* lineage are respiratory isolates vs. 101/213, 47% in all other isolates; Fisher’s exact p=0.0001), suggesting that this lineage is associated with increased capacity for respiratory colonization. Furthermore, a subset of isolates in the disrupted *kfoC* sub-lineage harbor the ICEKp10 element containing yersiniabactin. Examination of these genetic events in the context of the whole-genome phylogeny revealed that disruption of *kfoC* occurred first, followed by at least two different acquisitions of ICEKp10 (**Figures 4, S10**). Within the disrupted *kfoC* lineage, isolates with ICEKp10 are enriched in infection (31/55, 56% of isolates with ICEKp10 are infection isolates vs. 16/63, 25% of isolates without ICEKp10, Fisher’s exact p = 0.00065), supporting an increase in virulence after acquisition of ICEKp10. It is important to note that the observed clinical associations with ICEKp10 and kfoC disruption do not demonstrate causality, as we cannot rule out the role of correlated genetic variation.

**Figure 4:**
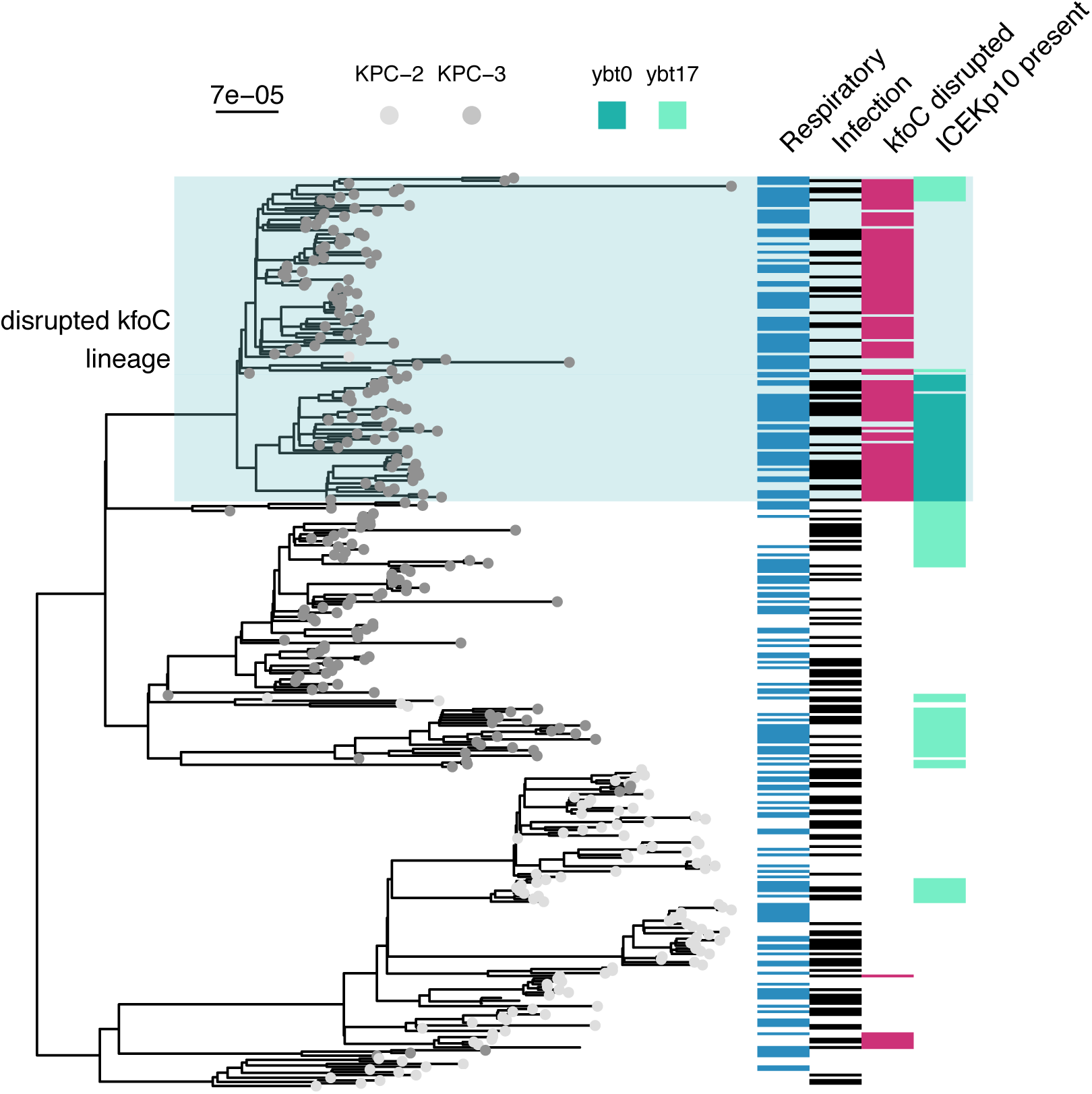
Select epidemiologic and genomic features visualized on the phylogeny indicate that a sub-clade of ST258 clade II may exhibit enhanced niche-specific adaptation and virulence. ST258 maximum likelihood phylogeny with the tip labels colored by KPC gene. The blue box indicates the sub-lineage with apparent altered niche-specific adaptation that acquires an additional virulence locus. The heatmap beside the tree indicates information about the isolate. From left to right: if it is a respiratory isolate, if it is an infection isolate, if *kfoC* is disrupted, and if it contains ICEKp10. Disrupted *kfoC* was associated with infection in the overall and respiratory machine learning analyses and ICEKp10 presence was associated with infection in the overall analysis. The scale bar to the top left of the phylogeny shows the branch length in substitutions per site. ybt=Yersiniabactin; ybt0 and ybt17 are two ybtSTs defined by Kleborate.

## Discussion

There have been numerous studies aimed at identifying risk factors for healthcare-associated infections caused by prominent antibiotic-resistance threats. For the most part, these studies have found the dominant risk factors to be linked to the magnitude of exposure (e.g. length of stay or colonization pressure), use of antibiotics, and overall comorbidity (43). Here we found similar results, where length of stay and having certain comorbidities were positively associated with infection. What remains unclear is, in the critically ill populations heavily exposed to antibiotics that are at greatest risk, if genetic variation in circulating resistant lineages influences patient infection status. Here, we addressed this question by focusing on CRKP infection in a comprehensively sampled cohort of patients from 21 LTACHs across the U.S. To gain a realistic assessment of the predictive capacities of patient and CRKP genetic features, we employed a machine learning framework using multiple facility-level train/test splits. Overall, we found that, while neither patient nor CRKP genetic features have high predictive accuracy on held-out test data, both feature sets were independently associated with infection, with one or the other being more predictive on different facility subsets. Moreover, the integration of clinical and genomic data led to the discovery of an emergent sub-lineage of the epidemic ST258 clone that may have increased adaptation for the respiratory tract, and is more strongly associated with infection.

One strength of our machine learning approach is that we were able to measure the variation in discriminative performance across 100 train/test iterations that differed in which facilities were included in train and test sets. We found that performance varied greatly depending on how facilities were allocated to train and test sets, highlighting how smaller studies could overestimate or underestimate the discriminative ability of both their model and individual features. Variation in model performance across facilities could be due to facility-level heterogeneity leading to differences in the prevalence of predictive patient or genomic features in the different test sets. For instance, certain facilities may have patient populations skewed towards individuals with characteristics that are predictive of infection. Alternatively, certain geographic regions may have CRKP strains that are more virulent than in other regions. These differences could lead to a higher predictive power for certain facilities compared to others. Another possible explanation for variation in model performance is that the critically-ill nature of LTACH patients may be such that most patients are actually highly susceptible to infection (i.e. many patients colonized with CRKP may ultimately develop an infection). However, it is noteworthy that despite these potential challenges in creating generalizable models, our analysis did yield predictors of infection and colonization consistent across test sets, and thus across LTACHs.

We built classifiers including all genomic features as well as a curated subset of features from Kleborate (16), and found that both are similarly weakly predictive of infection. However, while the uncurated feature set presented challenges with downstream interpretation, our analyses on the curated genomic features (16) facilitated novel insights into potential evolutionary trajectories of anatomic site-specific adaptation and virulence. For example, we observed that disruption of the O-antigen biosynthetic gene, *kfoC*, is associated with isolation from the respiratory tract. While we cannot determine from our machine learning analysis if disruption of *kfoC* is directly causal, the biological plausibility of an altered O-antigen structure mediating evasion of innate immunity and/or other beneficial interactions with the host makes this a strong candidate for follow-up experiments. Supporting this hypothesis, a previous study found that absence of O-antigen is associated with decreased virulence, but not decreased intrapulmonary proliferation, in a murine model (44). In addition, we noted that a number of antibiotic resistance determinants were associated with colonization. We hypothesize that this observation could be a consequence of longer duration of residence being associated with increased exposure to off-target antibiotics (45). Finally, we also saw evidence that, after acquiring the virulence factors yersiniabactin and colibactin on the ICEKp10 element, the disrupted *kfoC* subclade became more strongly associated with infection, supporting the idea that circulating ST258 sub-lineages can evolve to become both hypervirulent and multi-drug resistant (19, 46–48).

It is important to note that the machine learning method we employed does not correct for microbial population structure. We chose this method instead of alternative bacterial genome-wide association methods because our primary interest was in quantifying the overall predictive capacity of bacterial genotype in a patient population that was collected in a comprehensive and unbiased manner (i.e. all clinical isolates from 21 facilities over one year). While alternative methods controlling for population structure may yield more precise estimates of the contribution of individual variants, this would obfuscate the realized contribution in our patient population and hinder direct comparison to the predictive capacity of patient features. However, this then presented the challenge of interpreting our finding that certain sub-populations of CRKP may differ in their predilection for causing infections at different sites. For instance, the *kfoC* disruption is a lineage defining variant, and in principle other variants that define this lineage could also be causal. Here, we limited our analysis to a curated set of variants belonging to pathways known to be associated with antibiotic resistance and virulence, and found that only *kfoC* disruption was associated with increased respiratory infection, thus making it a strong candidate for follow-up in in vitro or in vivo models (10). To identify novel loci whose role in human infection may not be appreciated both computational and experimental strategies may be employed to help prioritize putative causal versus passenger variants. Computationally, investigators may search for evidence of parallelism in genotype/phenotype associations, which would bolster confidence in causality (49). Alternatively, high-throughput screens of genetic mutants in relevant model systems can help prioritize candidates. Garnering further genomic or experimental support for the direct role of a specific genetic variant would in turn increase the likelihood that those genetic markers would be predictive in new strains and patient populations.

Our study also has several important limitations related to the data available. Specifically, CRKP colonization vs. infection for non-bloodstream isolates may be difficult to discriminate using surveillance criteria and the clinical data that were available. However, we based our definitions on established CDC criteria with modifications used previously (7). Encouragingly, we were still able to identify consistent predictors of infection, even with potential misclassifications. A second limitation is that our dataset only included one clinical culture for the majority of patients, meaning that we were unable to investigate clinical or genomic features that may be associated with progression from colonization to infection. Furthermore, we do not have CRKP rectal colonization isolates, and therefore cannot evaluate transition from rectal colonization to other body sites. However, we hypothesize that comparing rectal colonization to infection may be asking a subtly distinct question - namely bacterial genetic factors that enable translocation from the gut to other body sites. In contrast, we hypothesize that our study design is ideal to identify bacterial genetic factors associated with infection once at a given body site. Additionally, we chose to focus our analysis on ST258 due to its disproportionate presence in our dataset, but this makes it possible that our findings may not generalize to other sequence types. Nevertheless, ST258 is the dominant clone in the U.S., and the methods we employed here can be used to study other sequence types and other pathogens. Furthermore, our focus on the ST258 lineage led to the particularly notable finding that even within an established endemic multi-drug resistant lineage (that emerged circa 2000 (14)), there is continued evolution that influences the manifestation and outcome of infection. We were also limited in the patient data included in our model. It is likely that important differences in underlying patient conditions were not captured by the coarse clinical variables we included, and we also did not account for differences in genetic variation in the host (50). Other limitations include that our study was restricted to LTACH patients, and had non-random geographic sampling. While LTACHs have unique structural features, based on prior studies, we expect that the types of patient risk factors considered are likely to generalize to other patient populations. Moreover, our restriction to LTACHs in endemic geographic regions has the benefit of focusing on populations at disproportionate risk for CRKP infection (8).

In conclusion, we employed a machine learning approach to quantify our ability to discriminate between CRKP colonization and infection using patient and microbial genomic features. This approach highlighted the high degree of variation in predictive accuracy across different facility subsets. Furthermore, despite modest predictive power, we identified several genomic features consistently associated with infection, indicating that variation in circulating CRKP strains contributes to infection, even in the context of the critically-ill patient populations residing in LTACHs. Future work should aim to corroborate our findings with larger cohorts, and follow up on strong associations to determine whether they are indeed risk factors for infection. This could ultimately help identify patients at high risk for infection and devise targeted strategies for infection prevention.

## Materials & Methods

### Clinical and genomic data

We used whole-genome sequences of clinical (non-surveillance) CRKP isolates and associated patient metadata from a prospective observational study performed in 21 LTACHs from across the U.S. over the course of a year (BioProject accession no. PRJNA415194) (14). We included only the first clinical bloodstream, respiratory, or urinary isolate from each patient (n=355; **Figure S1A**), and subset to only ST258 isolates for the majority of analyses (n=331; **Table S1**; see supplementary material for reasoning). Details about the analysis pipeline (15) genomic data curation (14, 16–22), and phylogenetic reconstruction (23–26) are provided in the supplementary material. While most clinical data cannot be shared, the deidentified patient ID, hospital of sample isolation, and isolation site are included in the Sequence Read Archive metadata for the BioProject.

### Outcome definition

Our outcome of interest was colonization vs. clinical infection (**Figure S1B**). Based on the U.S. Centers for Disease Control and Prevention’s (CDC) established National Healthcare Safety Network (NHSN) surveillance definitions, we considered all bloodstream isolates as representative of infection, and used modified definitions to classify urinary and respiratory cultures as representative of infection versus colonization (**Table S2**) (7, 27). Any isolate that did not meet the criteria for infection was classified as colonization.

### Feature sets

We studied the association between five different feature sets and infection/colonization in CRKP ST258 (**Figure S1C**); the feature sets are described below. See supplementary methods for details on feature set creation and processing. Counts below are for confident features from the entire dataset prior to subsetting for different analyses.

#### Patient

Clinical features described in Han *et al*. (n=50; **Table S3**) (14).

#### Uncurated genomic

Single nucleotide variants, indels, insertion sequence elements, and accessory genes (n=2447).

#### Uncurated grouped genomic

Variants grouped into genes (i.e. a burden test, e.g. (28)) and accessory genes (n=3159).

#### Curated genomic

Features identified by Kleborate (16), a tool designed to identify the presence of various genes and mutations known to be associated with either CRKP virulence or antibiotic resistance (n=91).

#### Patient & curated genomic

Patient features and curated genomic features (n=141).

### Machine learning & model selection

We aimed to classify clinical infection (vs. colonization) using each of the different feature sets (see above); we built classifiers using the first clinical isolate from each patient for all isolates, only respiratory isolates, and only urinary isolates. We performed L2 regularized logistic regression using a modified version of the machine learning pipeline presented in Topçuoğlu *et al*. (29) using caret version 6.0-85 (30) in R version 3.6.2 (31) (**Figure S1D1**). We randomly split the data into 100 unique ∼80/20 train/test splits, keeping all isolates from each LTACH grouped in either the training set or the held-out test set to control for facility-level differences among the isolates (e.g., background of circulating strains within each facility, patient population, and clinician test ordering frequency). For valid comparison, the train/test splits were identical across models generated with different feature sets. Hyperparameters were selected via cross-validation on the training set to maximize the average AUROC across cross-validation folds. See supplementary methods for more details.

### Model performance

We measured model performance using the median test area under the receiver operating characteristic curve (AUROC) and area under the precision recall curve (AUPRC), as well as the interquartile range, across all 100 train/test splits (**Figure S1D2**).

### Features consistently associated with colonization or infection

To determine the importance of each feature in predicting colonization vs. infection, we measured how much each feature influenced model performance by calculating feature importance using a permutation test (29) (**Figure S1D3**). For each combination of feature and data split, we randomly permuted the feature and calculated the ‘permuted test AUROC’ using the model generated with the training data. Features with a correlation of 1 were permuted together. We performed this permutation test 100 times for each feature/data split pair, and obtained a mean feature importance for each data split. A mean feature importance above zero indicates that that feature improved model performance for that data split. We highlight features where the mean permuted test AUROC was above zero in at least 75% of the data splits. In this way, the permutation importance method allows us to take into account the variation we observe across the 100 models, which is not possible with standard parametric statistical tests or odds ratios.

### Data analysis & visualization

See supplementary material for details on data analysis and visualization in R version 3.6.2 (31–37). All code and data that is not protected health information is on GitHub (https://github.com/Snitkin-Lab-Umich/ml-crkp-infection-manuscript).

## Data Availability

All code and data that is not protected health information is on GitHub.

https://github.com/Snitkin-Lab-Umich/ml-crkp-infection-manuscript

## Acknowledgements

We gratefully acknowledge Kindred Healthcare for their assistance in collecting data and isolates used in this study. We also thank Sean Muldoon, MD for his support and guidance throughout the study. Finally, we thank the patients and staff of the long-term acute-care hospitals (LTACHs) for their gracious participation in this study, Begüm Topçuoğlu for help with the machine learning pipeline, Ali Pirani for bioinformatics support, and members of the Snitkin lab, Mike Bachman, and Robert Weinstein for critical review of the manuscript.

## Authors’ contributions

ESS, JHH, EL, and ZL conceptualized the study and acquired funding to support the project. JHH and ZL performed data curation. ZL performed formal analysis, investigation, and visualization. ESS, JW, and ZL developed methodology. ESS supervised the project. ZL and ESS wrote the original draft. All authors reviewed and edited the manuscript.

## Supplementary materials

### Supplementary methods

#### Pipeline

We created a snakemake pipeline (1) to perform data pre-processing, machine learning analysis, and figure generation.

#### Genomic data

We used Kleborate version 0.3.0 (2) to identify sequence type, capsular types (K locus (3) and O locus (4)), virulence factor (5), and antibiotic resistance genes. In addition, we identified single nucleotide variants (SNVs) and indels by mapping reads to the KPNIH1 reference genome (BioProject accession number PRJNA73191; https://github.com/Snitkin-Lab-Umich/variant_calling_pipeline (6)). After variant calling, we identified the predicted functional impact of variants using SnpEff version 4.3T (7). We also identified large insertion sequence (IS) elements using panISa version 0.1.4 with the default settings (8), and accessory genome genes using Roary version 3.12.0 with the default settings (9).

#### Phylogenetic tree reconstruction

We constructed a phylogenetic tree of all ST258 isolates by first creating a whole-genome alignment by mapping reads to the KPNIH1 reference genome (GenBank accession number CP008827.1) using bwa mem version 0.7.12 (10); samtools version 1.9 (11) was used to generate binary alignment files. We then masked sites identified as recombinant by Gubbins version 2.3.2 (12) and used this masked whole-genome alignment to build a maximum likelihood phylogeny with IQ-TREE version 1.6.12 (13) using a GTR model of nucleotide substitution and ultra-fast bootstrap with 1000 replicates (-b 1000) (14). The tree was midpoint-rooted using the midpoint.root function in phytools version 0.6-99 (15), thus splitting the tree into ST258 clades I and II (16).

#### Phylogenetic clustering test

To determine whether infection isolates cluster on the phylogeny more than expected by random chance, we performed a permutation test (17). To identify the true number of isolates in a pure cluster while controlling for isolate clustering by LTACH, we enumerated isolates in pure infection or colonization subtrees that also contain isolates from more than one LTACH. Then we randomized the infection labels across isolates 1000 times and enumerated the number of isolates in a pure subtree for each randomization. We then compared the random counts of isolates in a pure cluster to the true number and calculated an empirical p-value.

#### Feature set pre-processing

We used five feature sets to study the association between infection and colonization. To determine how well machine learning models perform using a targeted vs. untargeted approach, we performed machine learning with uncurated genomic features as well as curated genomic features known to be related to virulence. In an attempt to increase power, we not only performed machine learning on individual uncurated genomic features, but also using a burden test where genomic variants were grouped into genes and features were presence/absence of a variant in a gene. This may help capture variation associated with infection at the gene level that might not be captured at the variant level (e.g. different samples can have different variants in the same gene that all lead to a similar function). For the uncurated grouped genomic analysis, grouped variants included SNVs and indels identified as moderate or high impact by SnpEff and IS elements in a gene or upstream of a gene. We also grouped modifier variants identified by SnpEff into intergenic regions. Only features that occurred in more than one isolate and less than n-1 isolates were included. For the uncurated genomic and uncurated grouped genomic datasets we collapsed genomic features with an identical pattern across all isolates to reduce machine learning runtime.

#### Machine learning rationale

We chose to perform L2 regularized logistic regression because it is easily interpretable, often performs just as well as more complicated methods when limited training data are available (18), and has a grouping effect, which means that all associated features are identified even if they are collinear (19).

#### Machine learning details

The machine learning pipeline removes variables with near-zero variance, normalizes all features to be between zero and one, and sets at least 20% of the data aside for testing in each train/test split. We split the training data into train and validate sets by LTACH using the groupKMultiFolds function from the caret R package (caret version 6.0-85) (20). Within each training set, we selected hyperparameters via five-fold cross-validation 100 times, maximizing the average cross-validation AUROC.

#### Comparing model performance between feature sets

To determine whether there was a significant difference in model performance between different feature sets, we calculated a two-sided empirical p-value for each identical train/test split using the formula (2 x min(fraction of AUROC differences ≥ 0, fraction of AUROC differences ≤ 0) (18).

#### Identification of IS element insertions in certain O2v2 kfoC genes

One of the curated genomic features we identified as associated with infection was what Kleborate called a missing gene (kfoC) in the O locus operon of certain O2v2 serotypes. On the Kleborate website, they indicate that a missing gene could represent a truly missing gene, or a gene that is split between two different contigs. To further investigate this, we used blastn version 2.9.0 (21) to search for kfoC genes in each of the samples. Additionally, we aligned the reads of all genomes to CP031810, a complete *K. pneumoniae* genome from PATRIC (22) using bwa mem version 0.7.12 (10), and samtools (11) was used to generate binary alignment files. Next, we used panISa (8) to identify IS element insertions in kfoC.

#### Data analysis & visualization

We used Fisher’s exact tests for bivariable analyses with categorical variables, and Wilcoxon rank-sum tests for bivariable analyses with continuous variables. We performed all data analysis and visualization in R version 3.6.2 (23) using the following packages: tidyverse version 1.3.0 (24), cowplot version 1.0.0 (25), ggtree version 2.0.1 (26, 27), ape version 5.3 (28), phytools version 0.6-99 (15), and grid version 3.6.2 (29).

## Supplementary results

### Patients with different sequence types show no substantive differences in infection status, anatomic site of isolation, or clinical characteristics

Over 90% of the isolates in our dataset were ST258 (**Table S2**), the dominant strain in the US (30). At the level of sequence type, we found no difference in infection prevalence (p=0.44) or anatomic site of isolation (p=0.66), indicating that at this coarse level there was not evidence of differences in strain virulence or adaptation to a certain anatomic site. Bivariable comparison of patient factors between patients with different sequence types revealed only four significant differences (unadjusted p-values < 0.05; previous use of piperacillin/tazobactam, cefepime, ciprofloxacin, or fluoroquinolones). As we found no substantive differences in infection, anatomic site of isolation, or patient variables when comparing different sequence types, we chose to focus all subsequent analyses on ST258, the dominant sequence type in our dataset. The genetic variation of isolates within ST258 is much smaller than the genetic variation between sequence types, so limiting our analyses to ST258 could improve our ability to identify associations of interest within ST258.

## Supplementary tables & figures

**Table S1:** Infection status and anatomic site of isolation by CRKP sequence type.

| Sequence Type | Colonization |  | Infection |  |  | Total (%) |
| --- | --- | --- | --- | --- | --- | --- |
|  | Respiratory | Urinary | Blood | Respiratory | Urinary |  |
| ST15 | 4 | 3 | 1 | 2 | 0 | 10 (2.8) |
| ST15-1LV | 0 | 0 | 0 | 1 | 0 | 1 (0.3) |
| ST17 | 0 | 0 | 0 | 2 | 0 | 2 (0.6) |
| ST2237 | 0 | 1 | 0 | 0 | 0 | 1 (0.3) |
| ST258 | 121 | 71 | 28 | 62 | 49 | 331 (93.2) |
| ST307 | 1 | 4 | 0 | 2 | 2 | 9 (2.5) |
| ST36 | 1 | 0 | 0 | 0 | 0 | 1 (0.3) |

**Table S2:** Definitions of infection for urinary and respiratory cultures.

| Source | Infection definition |
| --- | --- |
| Urinary | ≥100,000 colony-forming units/mL on urinary culture with no more than 2 species of microorganisms AND at least one of the following:<br>• Leukocytosis (≥10,000 cells/μL) ≤ 7 days prior to specimen collection<br>• Positive urinalysis demonstrated by at least one of the following findings:<br>○ Positive dipstick for leukocyte esterase and/or nitrate<br>○ Pyuria (urinary specimen with ≥ 10 white blood cells/mL or ≥ 3 white blood cells/high power field of unspun urinary)<br>○ Microorganisms seen on Gram stain of unspun urinary |
| Respiratory | Chest X-ray with interpretation of pneumonia, probable pneumonia, or infiltrate/consolidation AND leukocytosis ($\geq 10,000$ cells/$\mu\text{L}$) $\leq 7$ days prior to specimen collection |

**Table S3:**
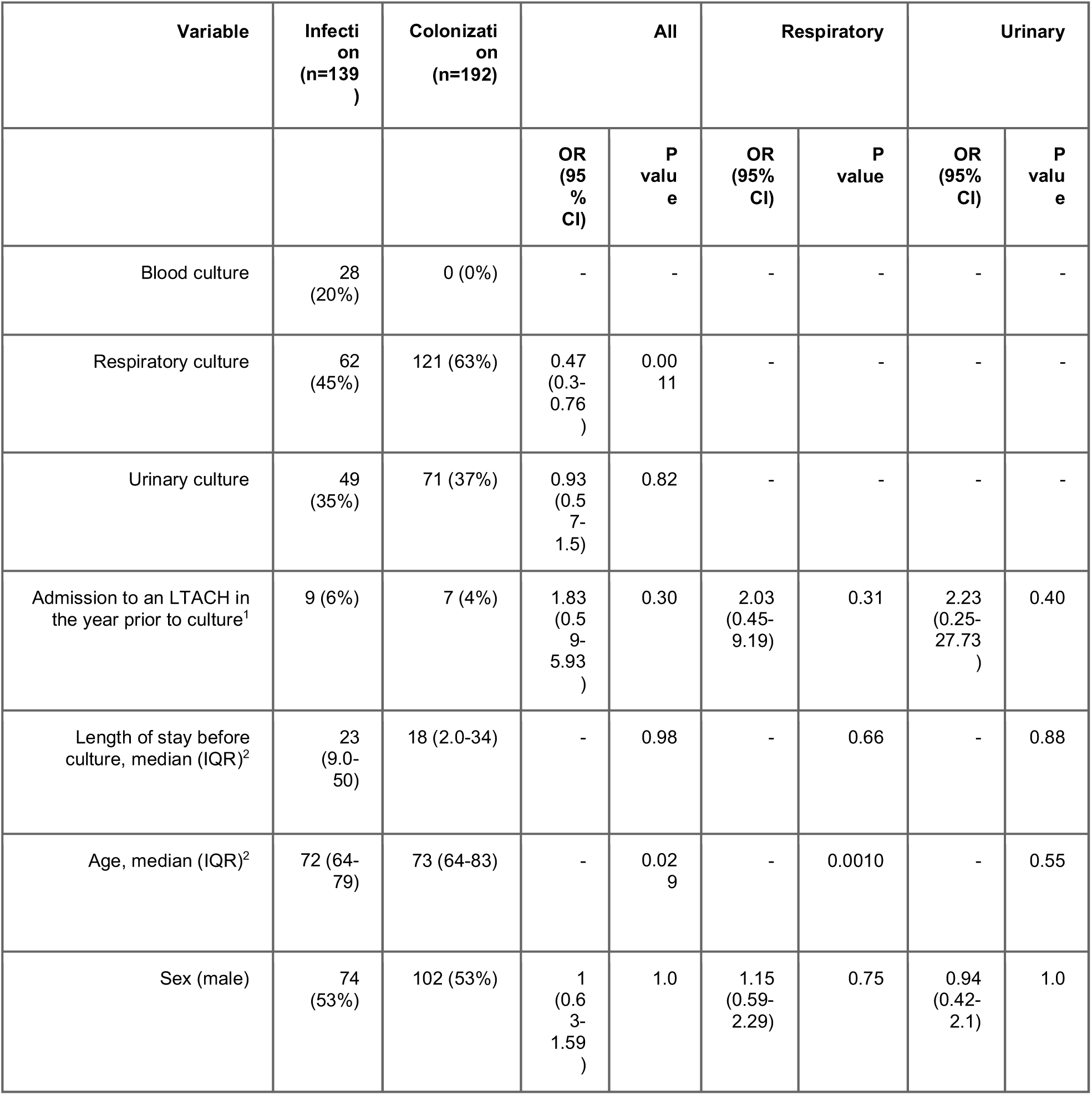
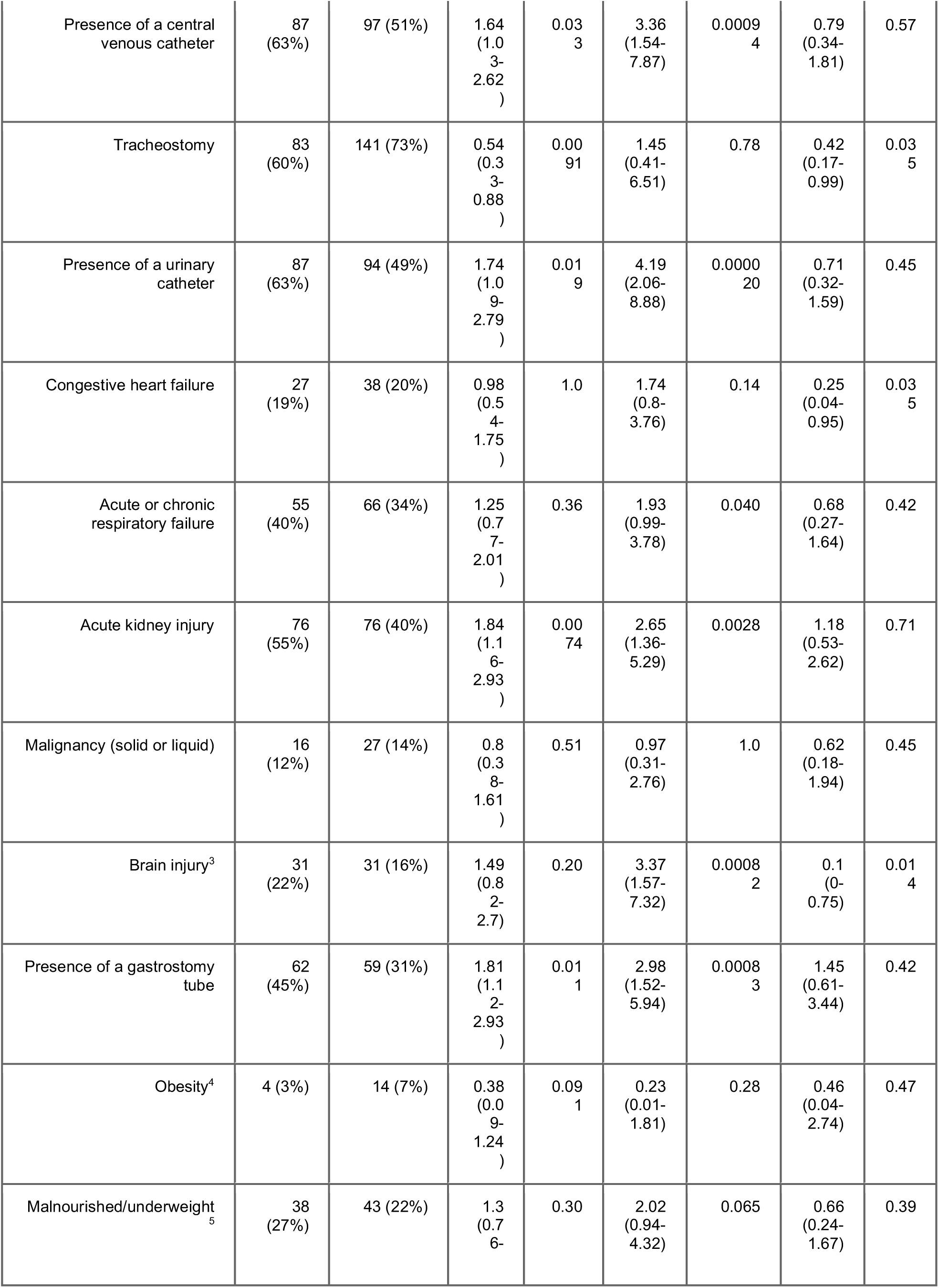
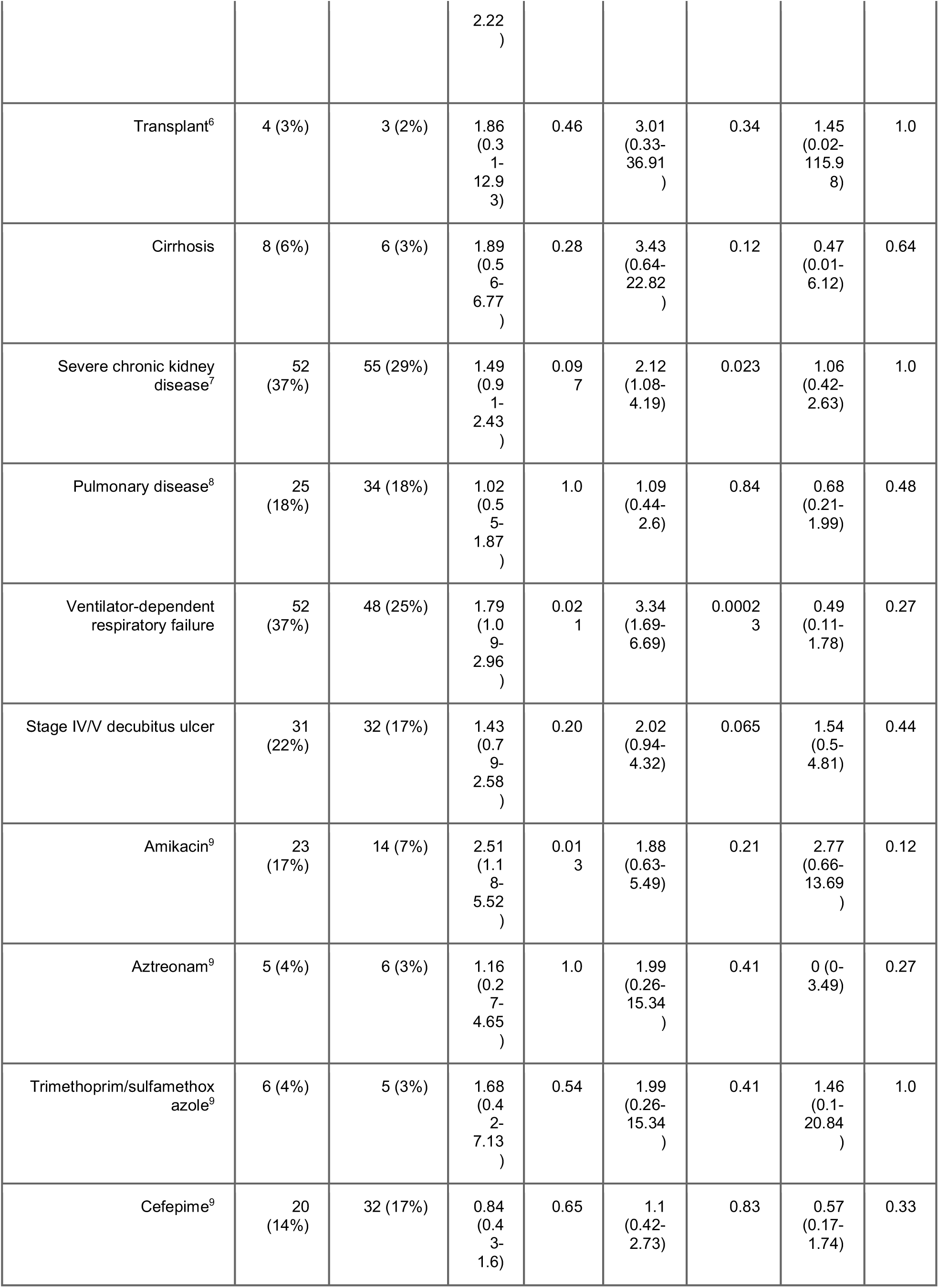
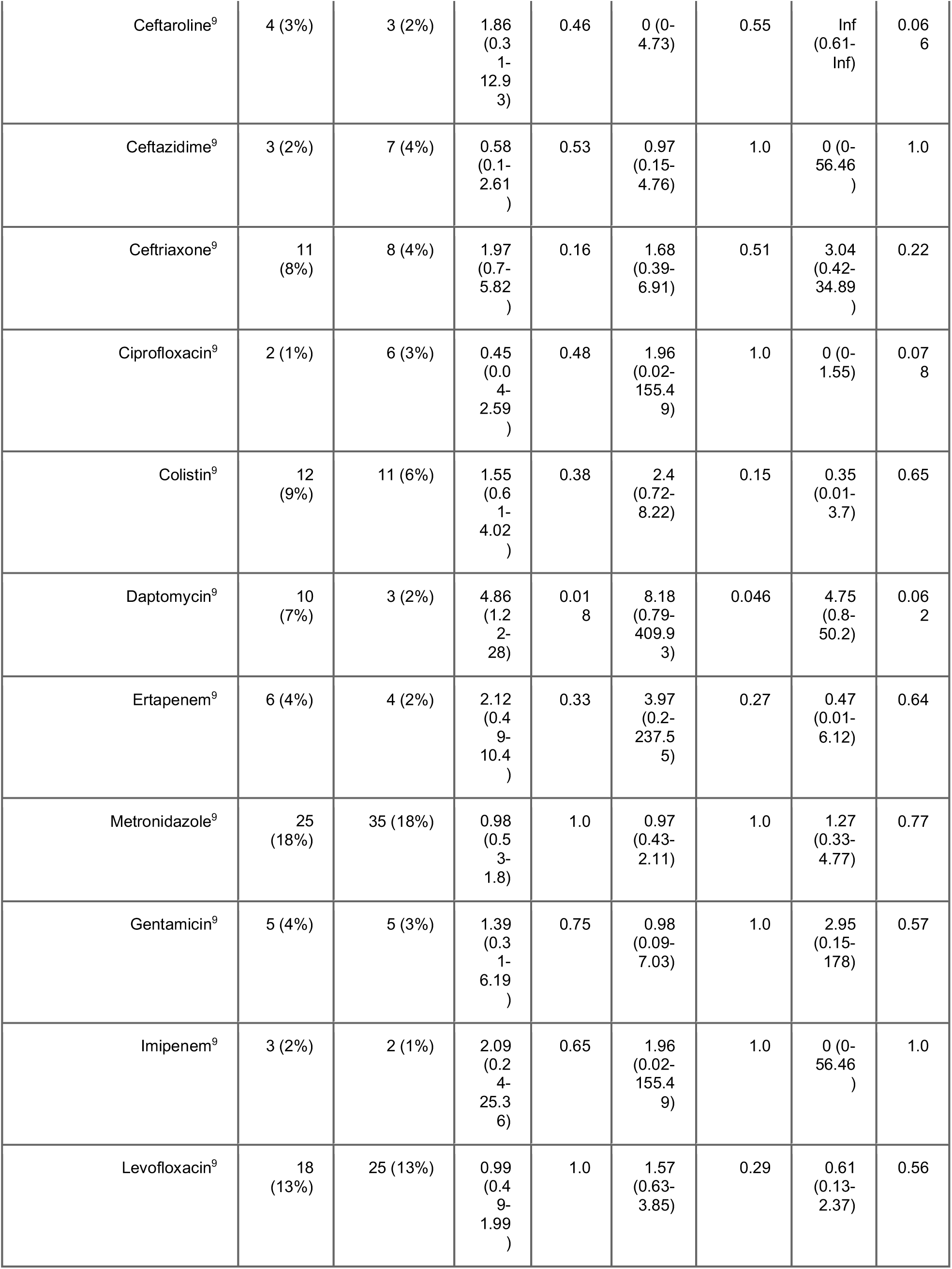
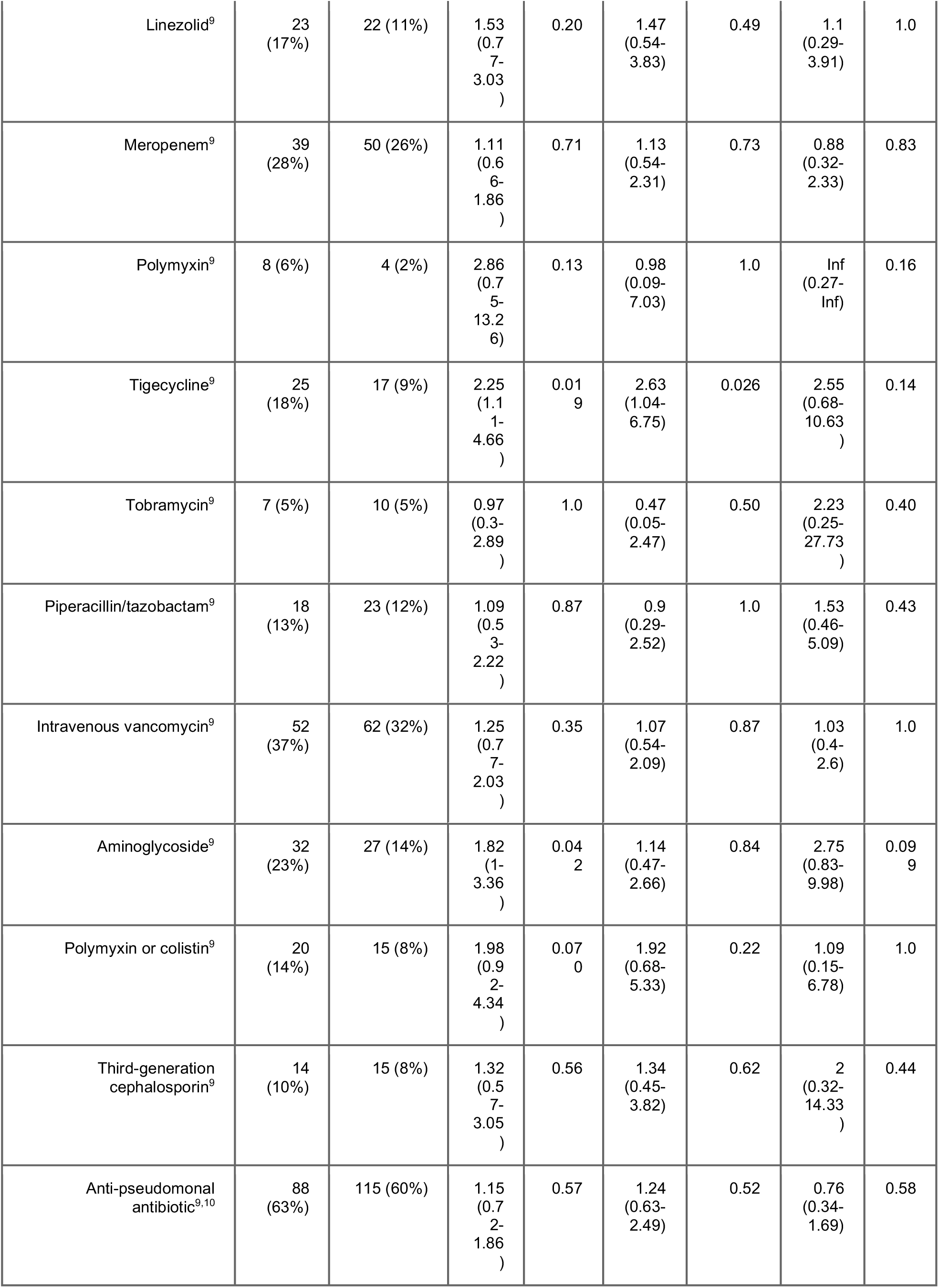
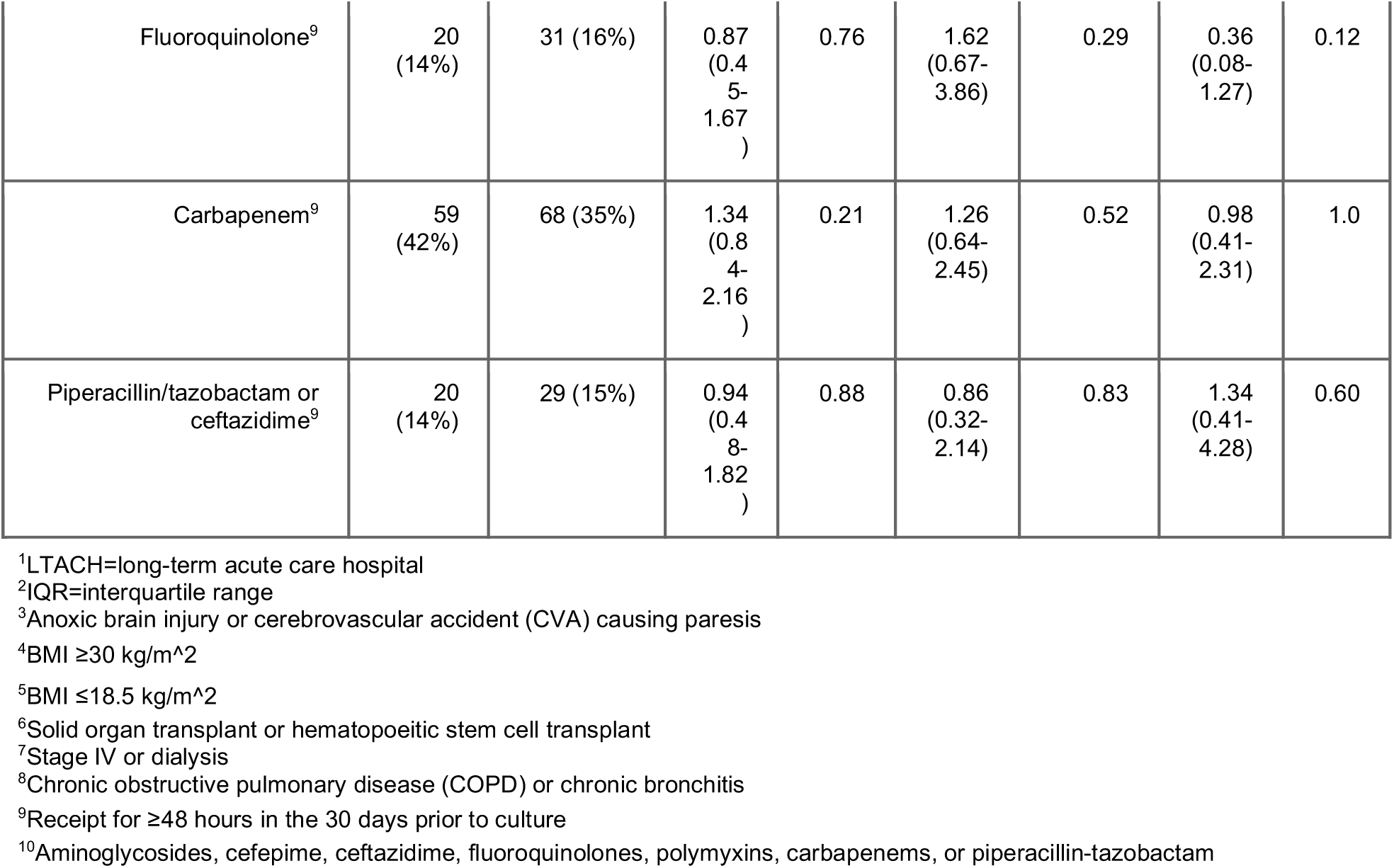
Bivariable analysis of patient features associated with CRKP ST258 infection in LTACH residents.

**Figure S1:**
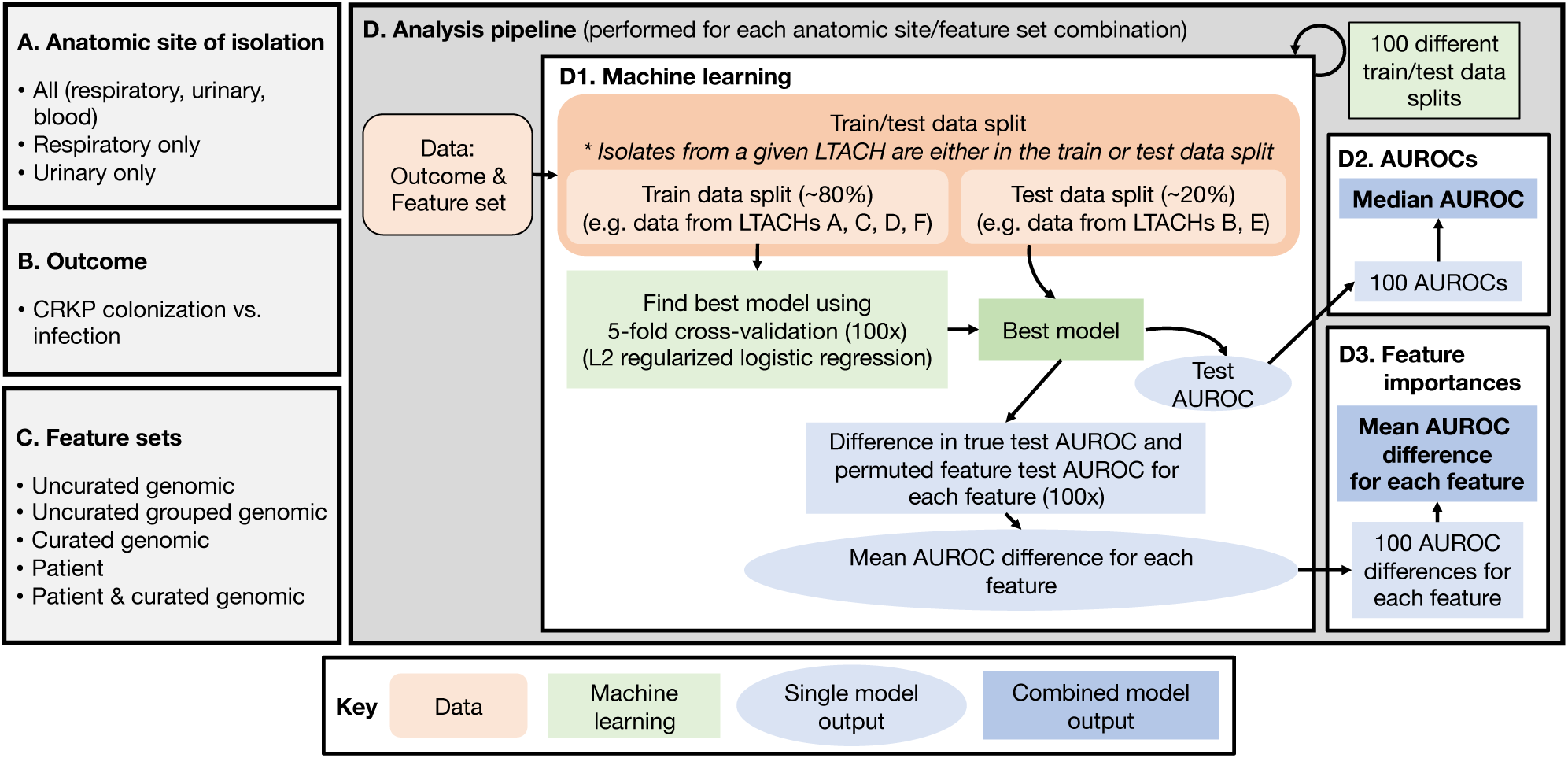
Methods overview. (A) Isolate subsets, based on anatomic site, used for different machine learning analyses. (B) Outcome used for all machine learning analyses. (C) Feature sets used for machine learning analyses (see main methods for more details). (D) Machine learning analysis pipeline: (D1) Machine learning pipeline, (D2) Median AUROC calculated from machine learning pipeline, (D3) Mean difference in test vs. permuted AUROC for each feature (see methods for more details). AUROC=area under the receiver operating characteristic curve.

**Figure S2:**
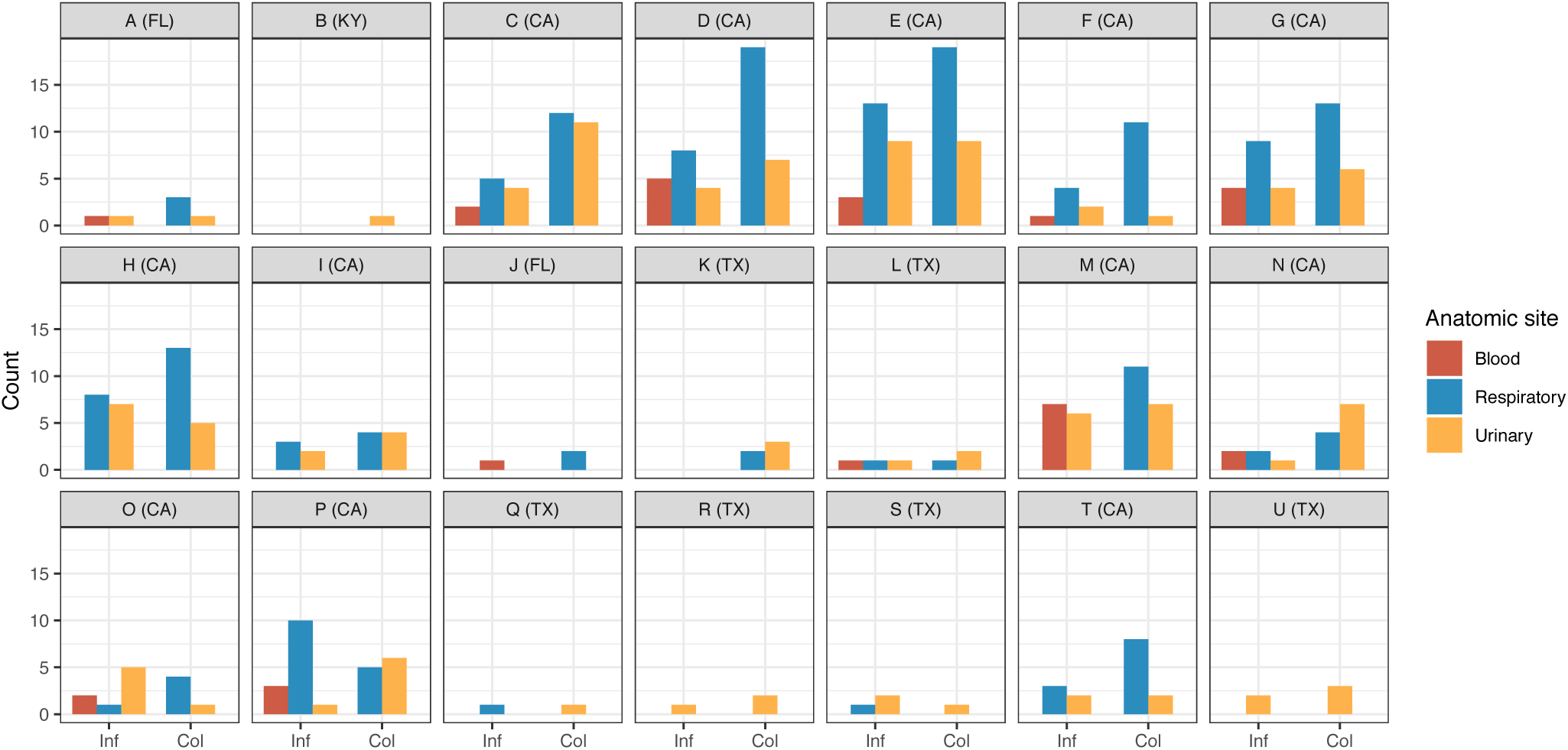
There are differences in sample size and distribution of infection and anatomic site of samples across LTACHs. Distribution of colonization and infection isolates are split by source and LTACH of origin. A to U are specific LTACHs and the geographic location by state is in parentheses. LTACH=long-term acute care hospital.

**Figure S3:**
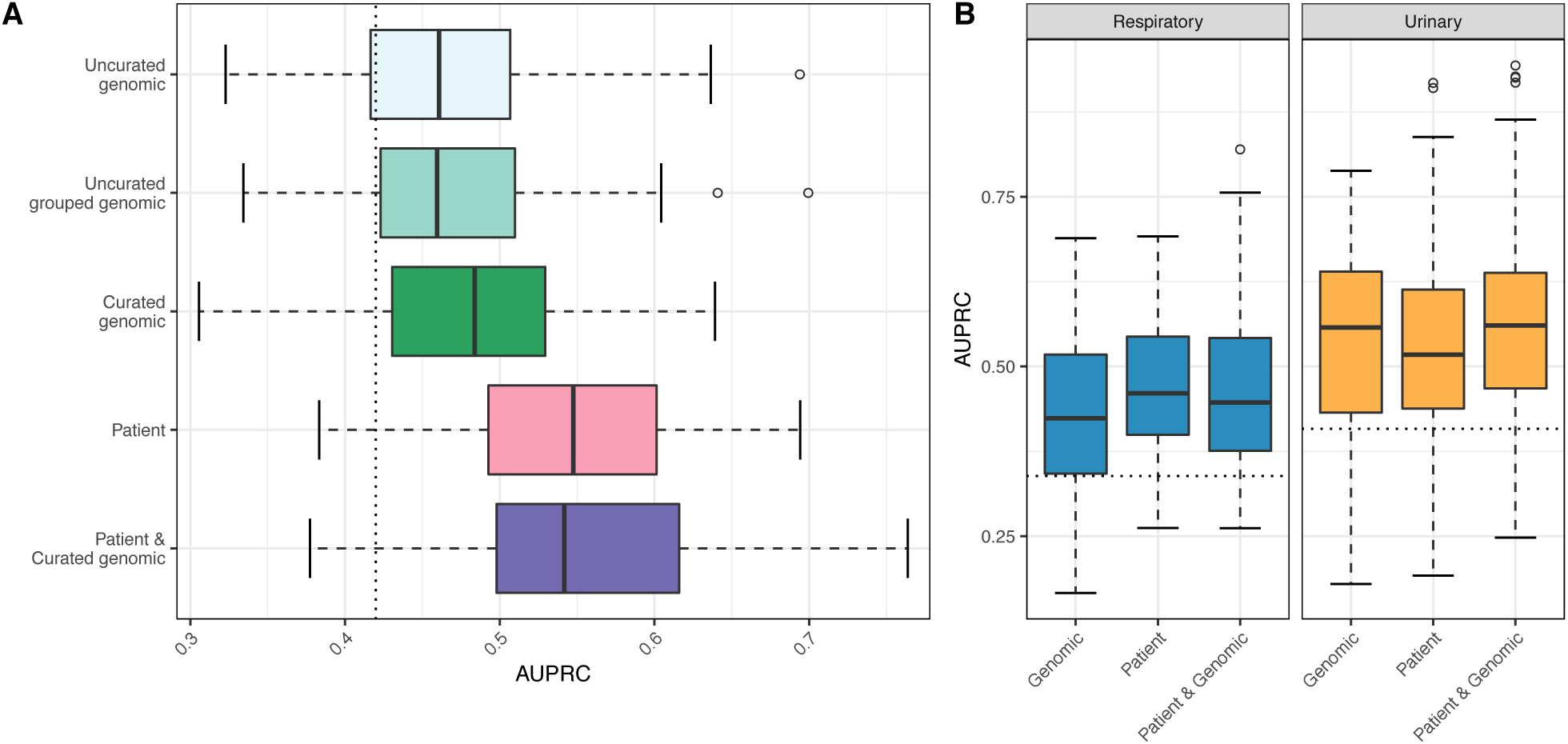
Test AUPRCs for various classifiers identifying CRKP colonization vs. infection for the overall machine learning model and respiratory and urinary models. Test AUPRCs for 100 train/test splits used to build classifiers with L2 regularized logistic regression. (A) Overall model with various classifiers. (B) Respiratory and urinary models; the curated genomic feature set was used here AUPRC=area under the precision recall curve.

**Figure S4:**
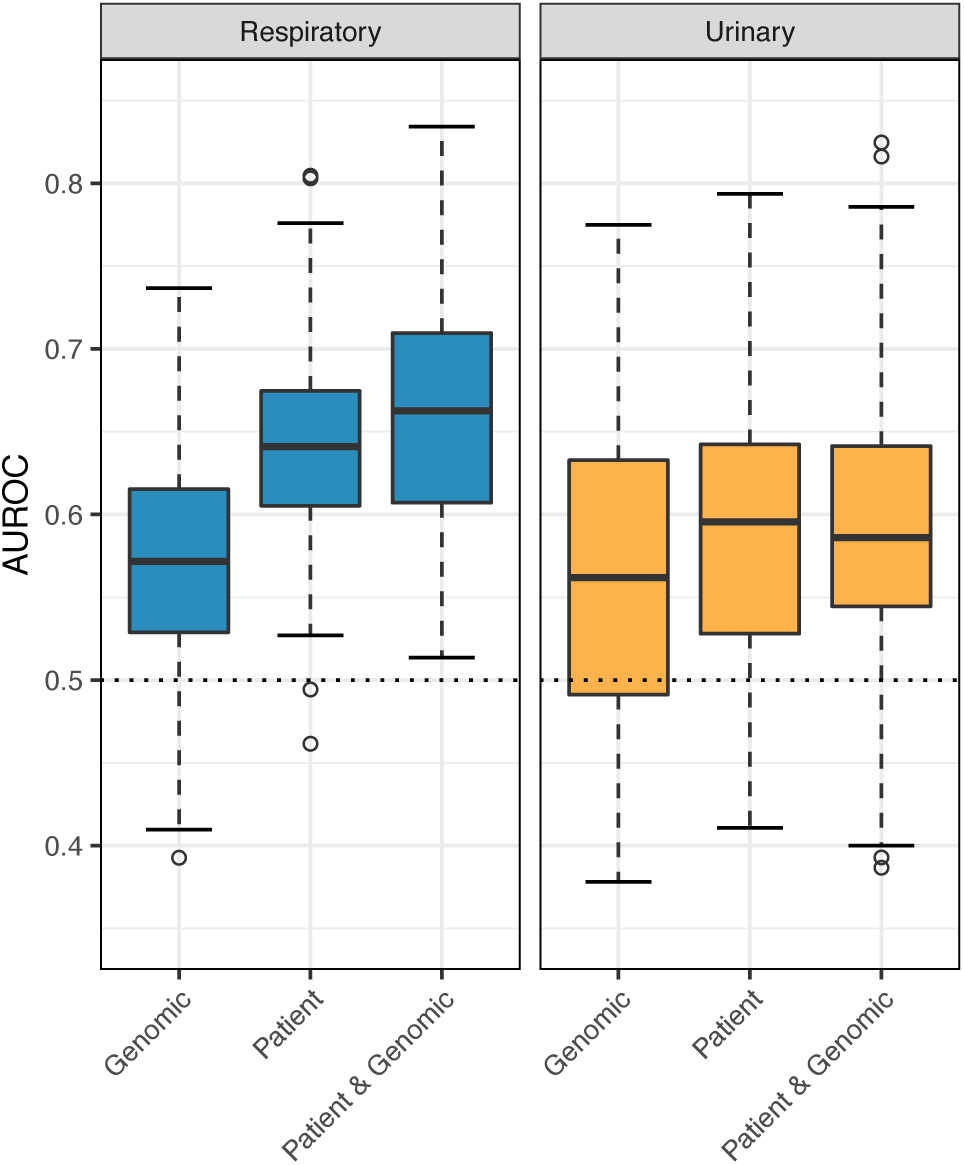
Test AUROC of respiratory and urinary ST258 only machine learning models for various feature sets. The curated genomic feature set was used here. AUROC=area under the receiver operating characteristic curve.

**Figure S5:**
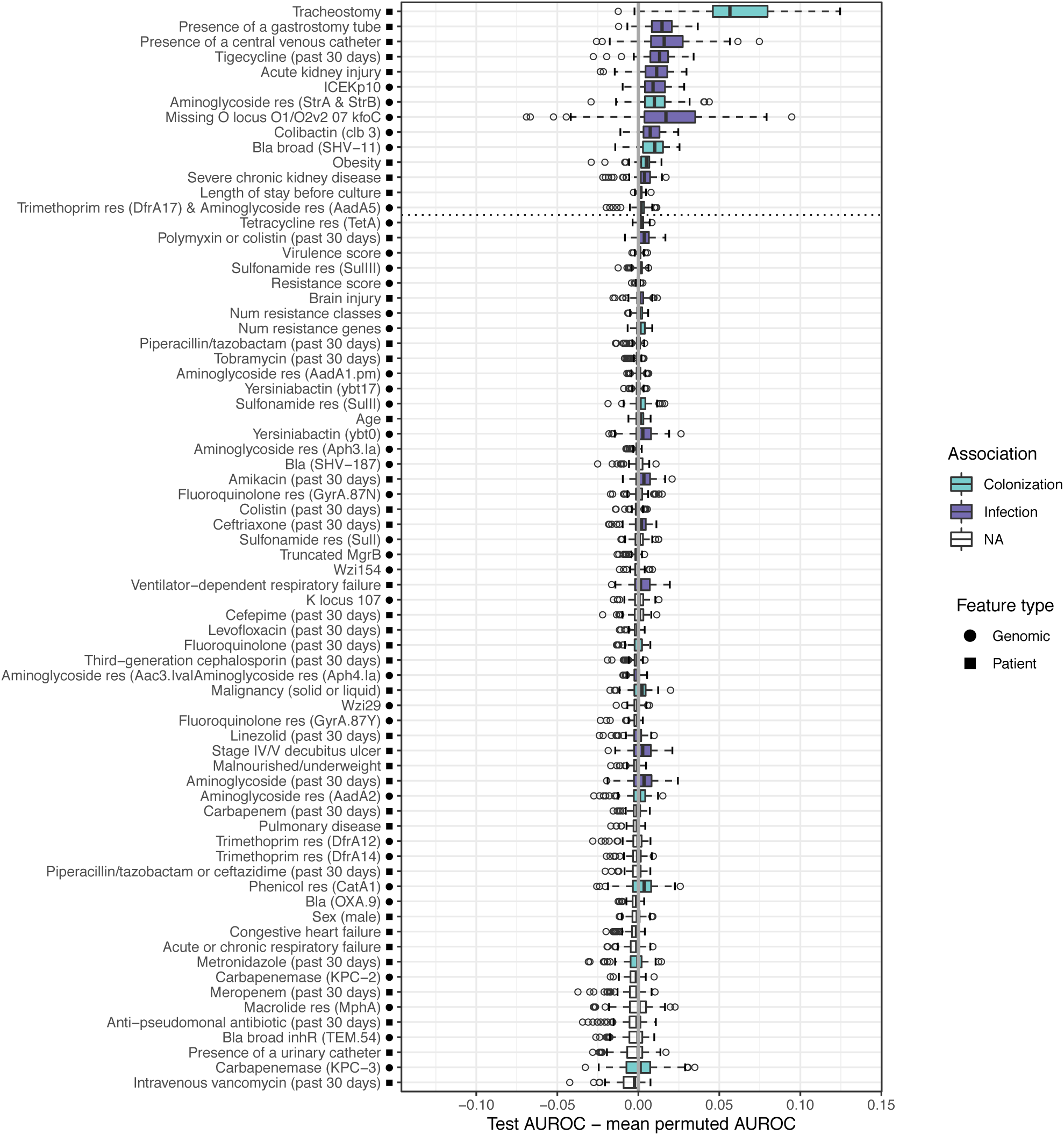
Differences in test vs. mean permuted AUROC across 100 data splits for each feature in the overall machine learning analysis. All features included in the model are shown. We consider features to be associated with infection/colonization if the AUROC difference is greater than zero in over 75% of the 100 data splits (see methods for more details). Features above the dotted line fall into this category. The solid grey line indicates a difference of zero (i.e. the feature provides no improvement to model performance). Bla=Beta lactamase, res=confers resistance to that antibiotic class.

**Figure S6:**
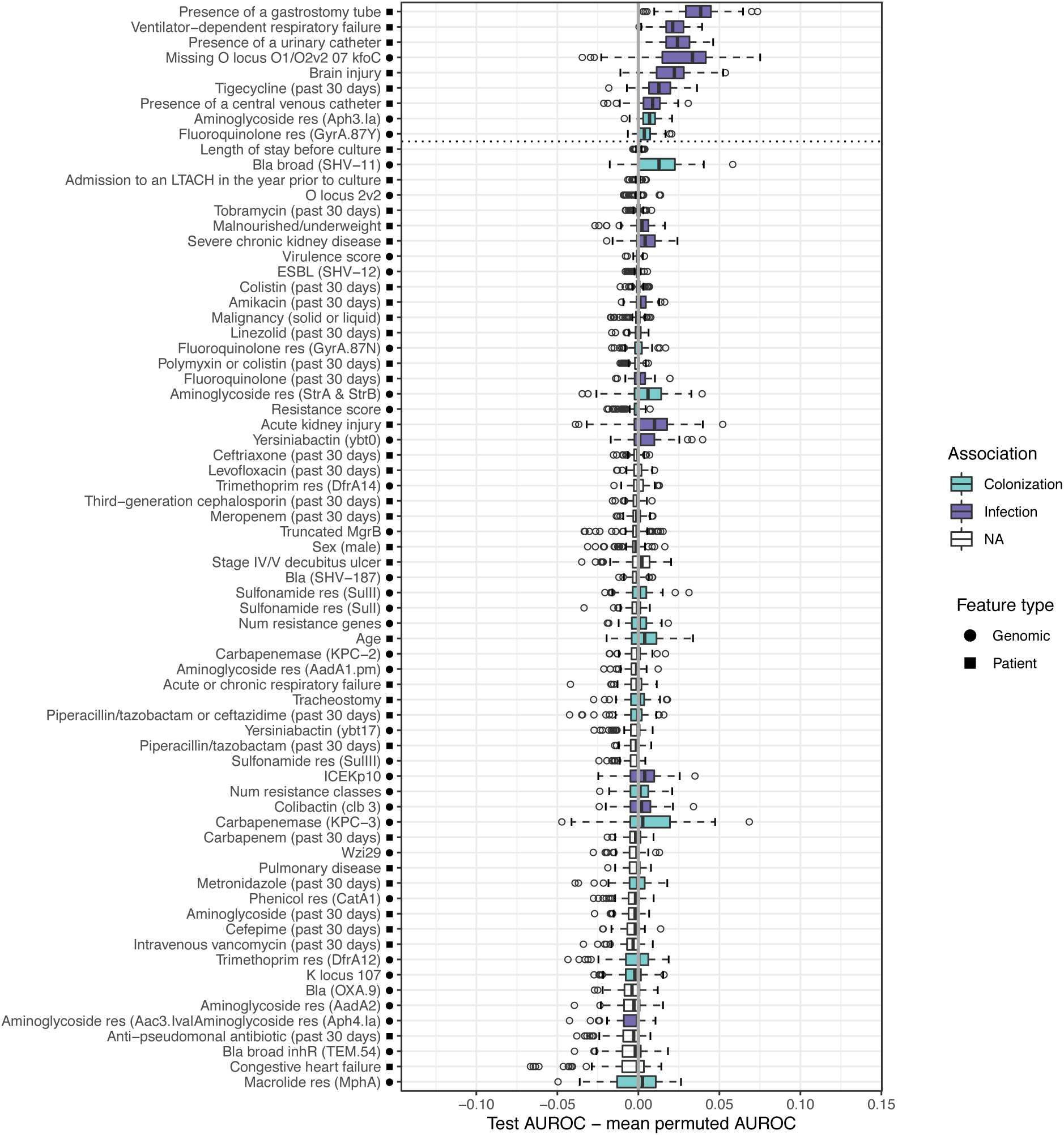
Differences in test vs. mean permuted AUROC across 100 data splits for each feature in the respiratory machine learning analysis. All features included in the model are shown. We consider features to be associated with infection/colonization if the AUROC difference is greater than zero in over 75% of the 100 data splits (see methods for more details). Features above the dotted line fall into this category. The solid grey line indicates a difference of zero (i.e. the feature provides no improvement to model performance). Bla=Beta lactamase, res=confers resistance to that antibiotic class.

**Figure S7:**
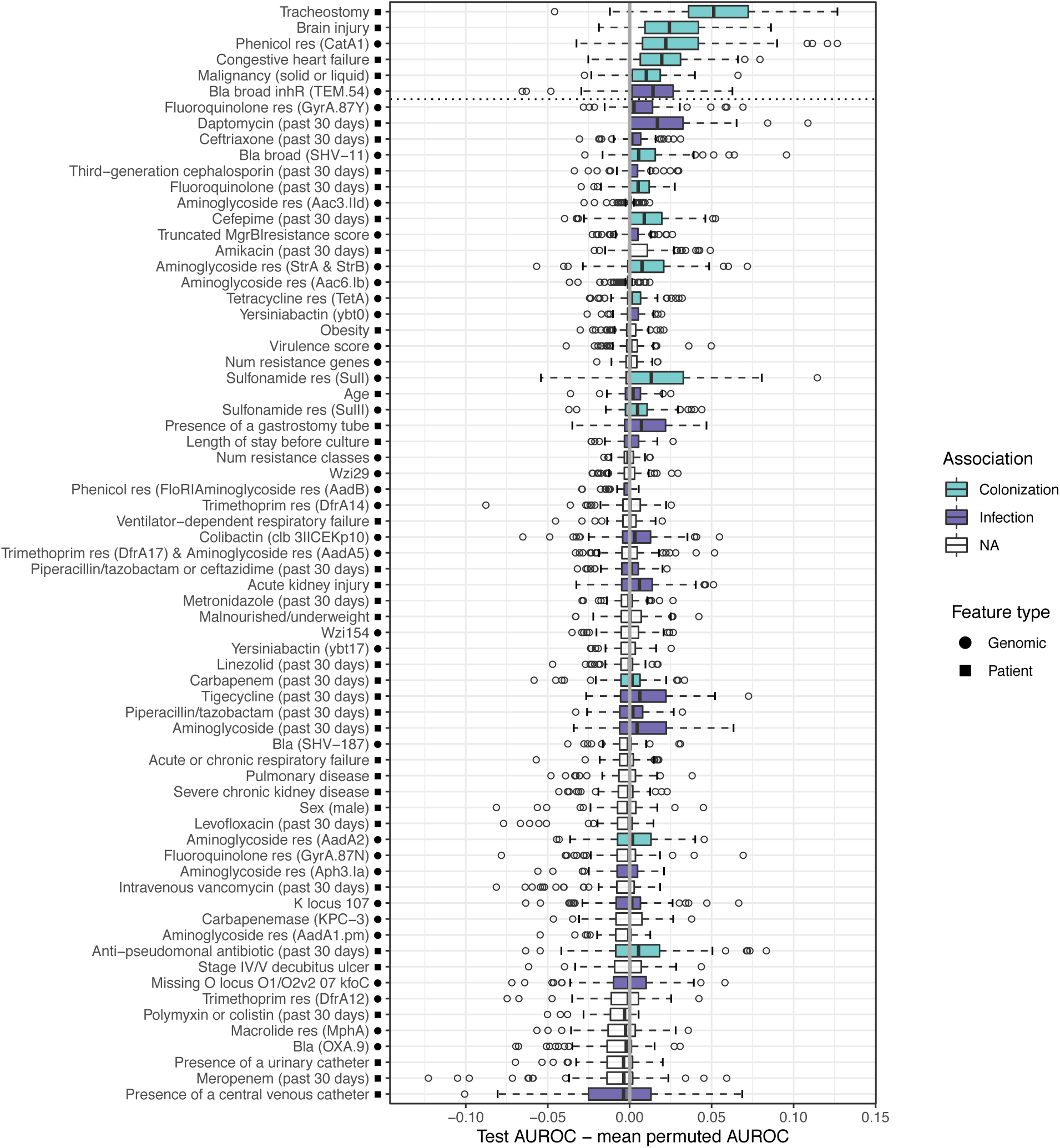
Differences in test vs. mean permuted AUROC across 100 data splits for each feature in the urinary machine learning analysis. All features included in the model are shown. We consider features to be associated with infection/colonization if the AUROC difference is greater than zero in over 75% of the 100 data splits (see methods for more details). Features above the dotted line fall into this category. The solid grey line indicates a difference of zero (i.e. the feature provides no improvement to model performance). Bla=Beta lactamase, res=confers resistance to that antibiotic class.

**Figure S8:**
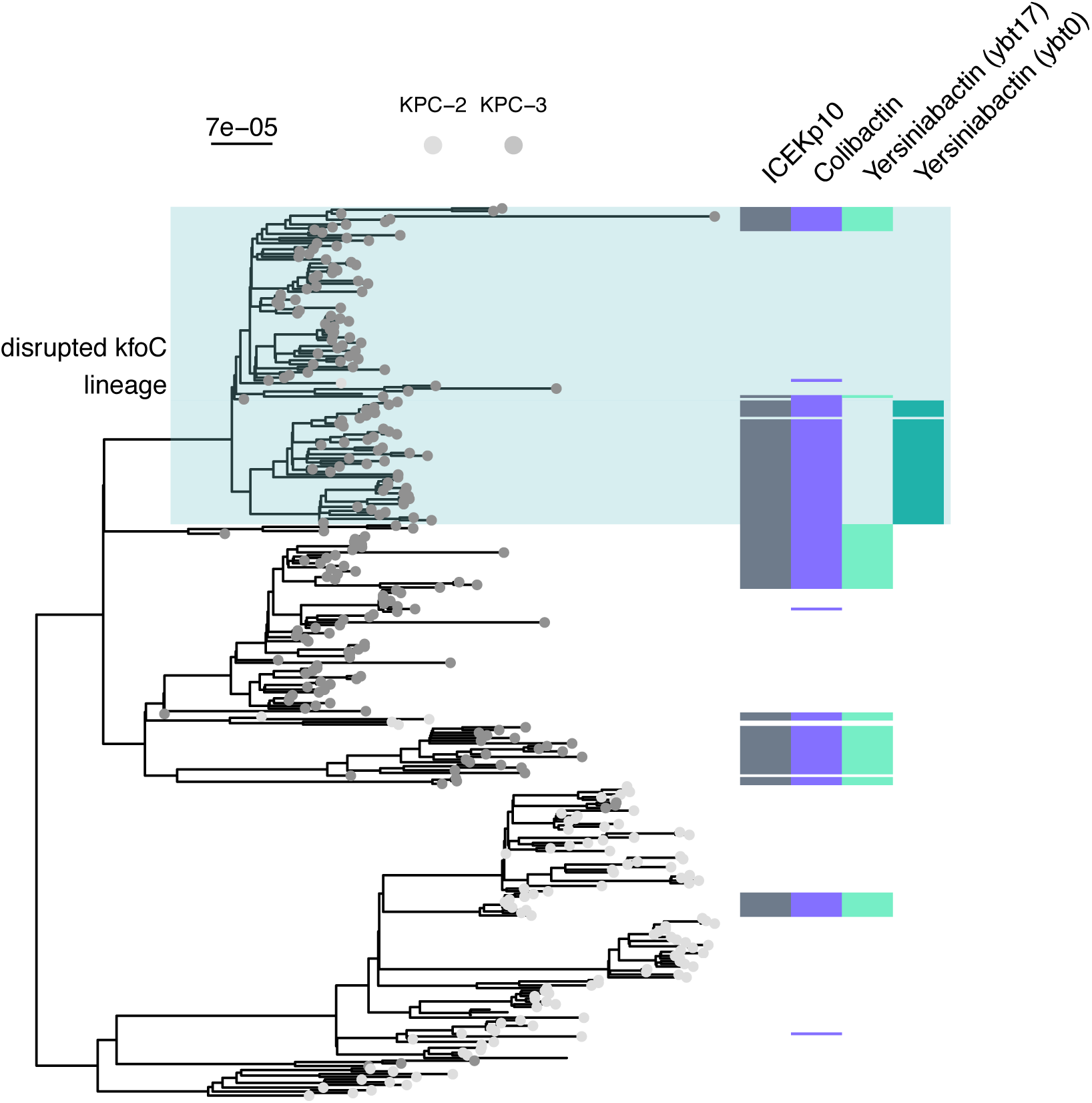
Presence/absence of genomic variants on the ICEKp10 element. The heatmap beside the ST258 phylogeny indicates variant presence/absence in a given isolate. Color indicates presence and white indicates absence. The scale bar above the phylogeny shows the branch length in substitutions per site.

**Figure S9:**
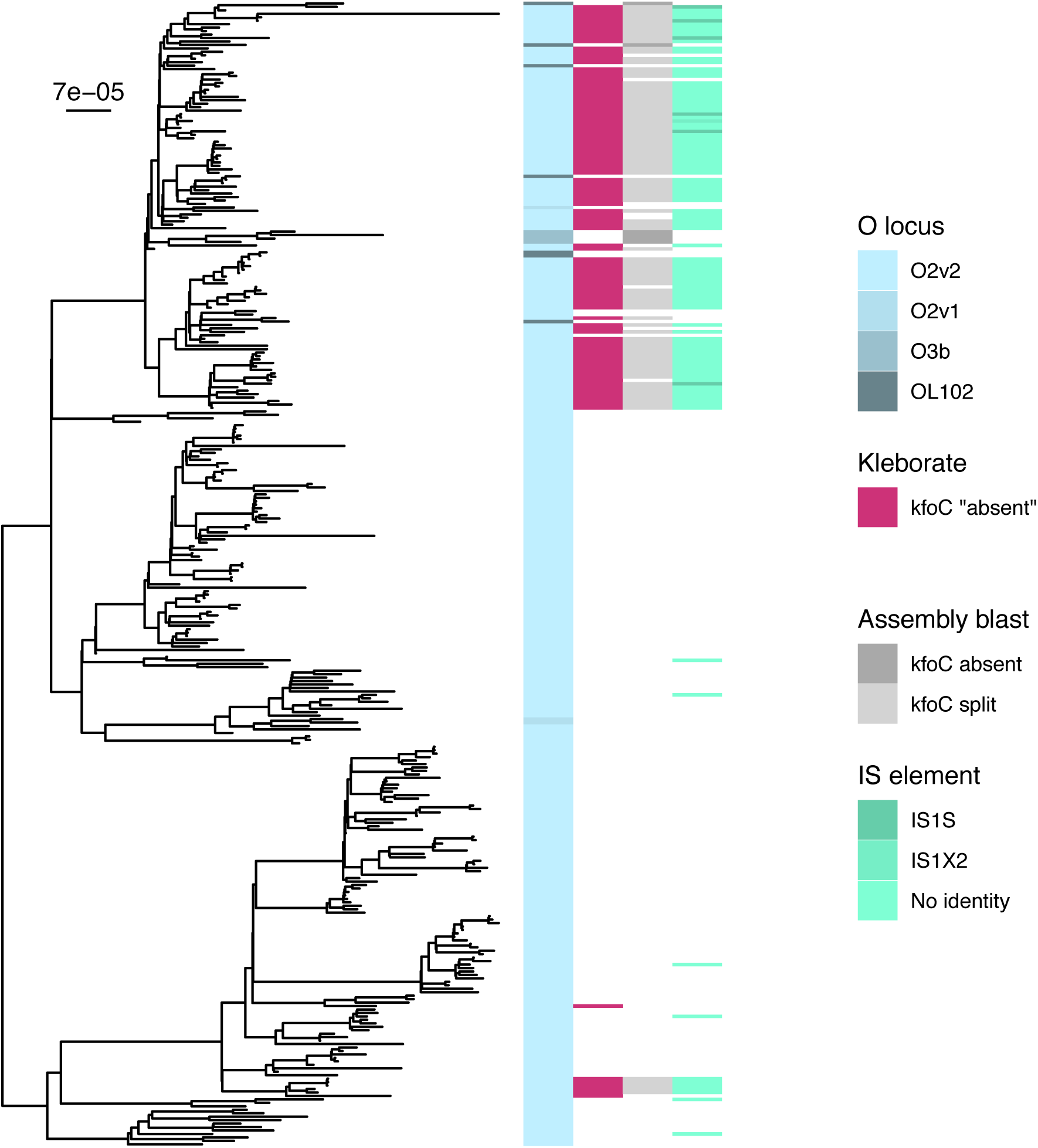
kfoC contains an IS element in a subset of ST258 isolates. The heatmap beside the ST258 phylogeny indicates information about the isolate (left to right): the O locus type identified by Kleborate (i.e. the curated genomic data), if Kleborate identified kfoC as missing, blast results for kfoC against the genome assemblies (split between 2 assemblies or entirely absent; white indicates that kfoC was intact), and IS elements identified in kfoC (white indicates no IS element detected). The scale bar to the top left of the phylogeny shows the branch length in substitutions per site.

**Figure S10:**
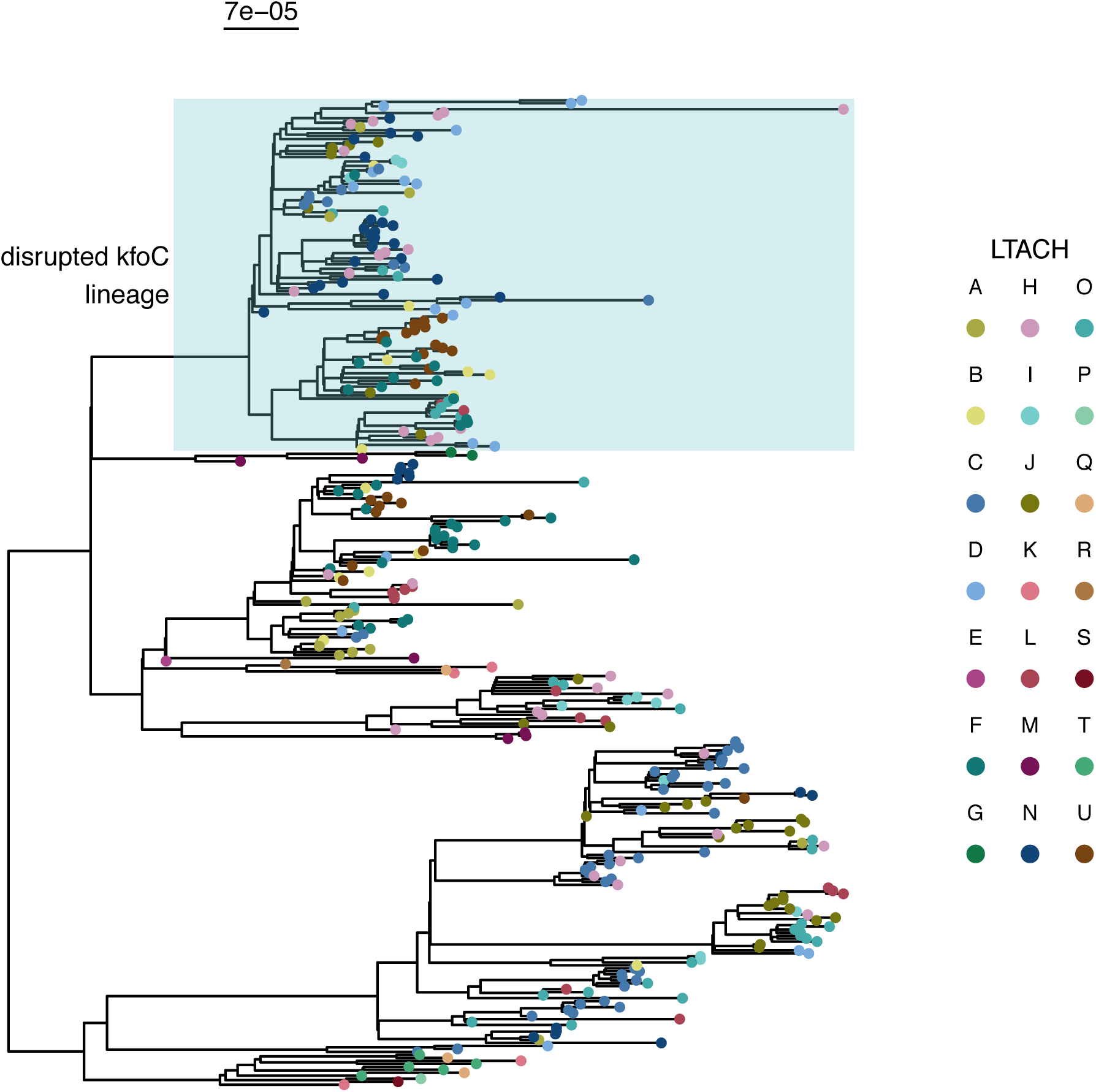
Isolates from different LTACHs are distributed across the phylogeny. Phylogeny of isolates indicating the LTACH the patient was in when the isolate was obtained. Isolates from several LTACHs are present in the disrupted kfoC lineage or the ybt0 lineage, and isolates from these LTACHs are not confined to those lineages. The scale bar above the phylogeny shows the branch length in substitutions per site.

**Figure S11:**
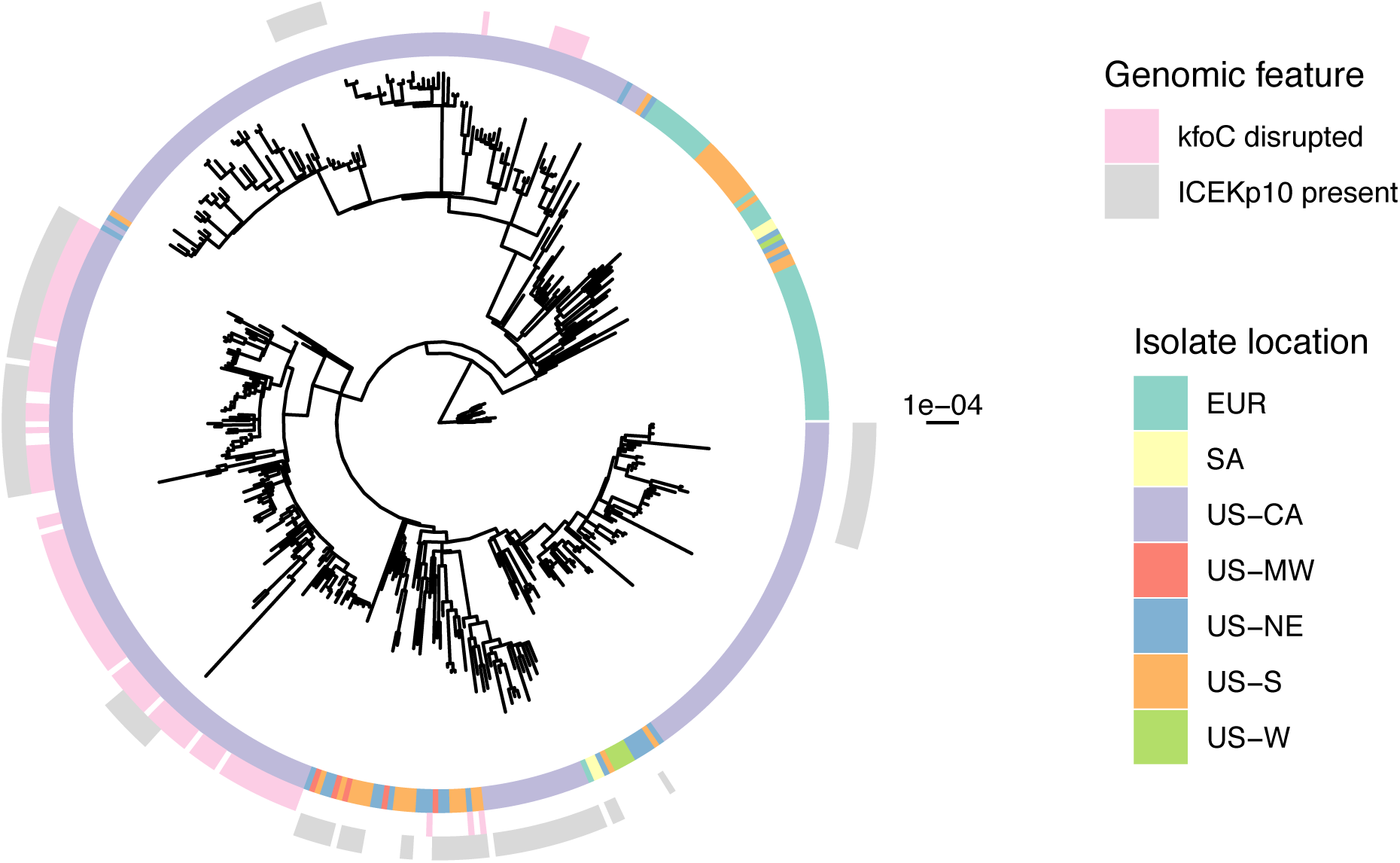
Isolates containing both disrupted kfoC and ICEKp10 are mainly confined to one ST258 clade. Phylogeny of California LTACH isolates (US-CA) in the context of public isolates from various locations (EUR=Europe, SA=South America, US-MW=US Midwest, US-NE=US Northeast, US-S=US South, US-W=US West). Isolate location, as well as ybt0 presence and kfoC absence are displayed for each sample. The scale bar to the right of the phylogeny shows the branch length in substitutions per site.

